# COVID-19: Risks of Re-emergence, Re-infection, and Control Measures – A Long Term Modeling Study

**DOI:** 10.1101/2020.09.19.20198051

**Authors:** Subhas Kumar Ghosh, Sachchit Ghosh

## Abstract

In this work we define a modified SEIR model that accounts for the spread of infection during the latent period, infections from asymptomatic or pauci-symptomatic infected individuals, potential loss of acquired immunity, people’s increasing awareness of social distancing and the use of vaccination as well as non-pharmaceutical interventions like social confinement. We estimate model parameters in three different scenarios - in Italy, where there is a growing number of cases and re-emergence of the epidemic, in India, where there are significant number of cases post confinement period and in Victoria, Australia where a re-emergence has been controlled with severe social confinement program. Our result shows the benefit of long term confinement of 50% or above population and extensive testing. With respect to loss of acquired immunity, our model suggests higher impact for Italy. We also show that a reasonably effective vaccine with mass vaccination program can be successful in significantly controlling the size of infected population. We show that for India, a reduction in contact rate by 50% compared to a reduction of 10% in the current stage can reduce death from 0.0268% to 0.0141% of population. Similarly, for Italy we show that reducing contact rate by half can reduce a potential peak infection of 15% population to less than 1.5% of population, and potential deaths from 0.48% to 0.04%. With respect to vaccination, we show that even a 75% efficient vaccine administered to 50% population can reduce the peak number of infected population by nearly 50% in Italy. Similarly, for India, a 0.056% of population would die without vaccination, while 93.75% efficient vaccine given to 30% population would bring this down to 0.036% of population, and 93.75% efficient vaccine given to 70% population would bring this down to 0.034%.

## Introduction

In December 2019, an outbreak occurred in Wuhan, China involving a zoonotic coronavirus, similar to the SARS coronavirus and MERS coronavirus^1^. Subsequently, the virus has been named Severe Acute Respiratory Syndrome Coronavirus 2 (SARS-CoV-2), and the disease caused by the virus has been named the coronavirus disease 2019 (COVID-19). Since then the ongoing pandemic has infected more than 30 million people and has caused more than 900 thousand deaths worldwide.

Patients with SARS-CoV-2 infections have mild to severe respiratory illness with symptoms such as fever, cough and shortness of breath. For majority of the patients, these symptoms appear 2–14 days after exposure, and for majority the symptoms are not life threatening. However, it has been reported that there are patients who are diagnosed by a positive RT–PCR test but are either asymptomatic or minimally symptomatic^2–6^. There are reasonable evidence to consider that such asymptomatic or minimally symptomatic individuals have longer duration of viral shedding than the symptomatic individuals and can spread the virus to susceptible group^6^. Hence, it is possible that the spread of SARS-CoV-2 is much higher and such undetermined transmission has been playing an important role in sustaining the community spread.

Mathematical Modeling plays an important role in understanding the trajectory of epidemic and design effective control measures under set of assumptions^7–9^. Here, we propose a new deterministic compartmental model for the COVID-19 epidemic that extends the classical SEIR (susceptible, exposed, infectious, recovered) model. We define the model and its parameters to address three different scenarios -in Italy, where there is a growing number of cases and re-emergence of the epidemic, in India, where there are significant number of cases post confinement period and in Victoria, Australia where a re-emergence has been controlled with severe social confinement program. In our model we also consider long term scenarios including re-emergence, re-infection, and control measures like mass vaccination program. Other than simulating these scenarios, another important distinction of our model is that we use parametric functions for contact rate, testing and vaccination.

We use a deterministic compartmental model that is an extension of the SEIR model^8,10^ in which we include current experience with SARS-CoV-2. We partition the total population into susceptible individuals (*S*(*t*)), exposed individuals (*E*(*t*)), Asymptomatic, undetected and infected individuals (*A*(*t*)), Symptomatic, undetected, and infected individuals (*I*(*t*)), Asymptomatic, diagnosed and infected individuals (*Q*(*t*)), Symptomatic, diagnosed, and infected individuals (*H*(*t*)), individuals with acute symptoms and in critical care (*C*(*t*)), and recovered (*R*(*t*)) and deceased (*D*(*t*)), see Figure 1.

**Figure 1.**
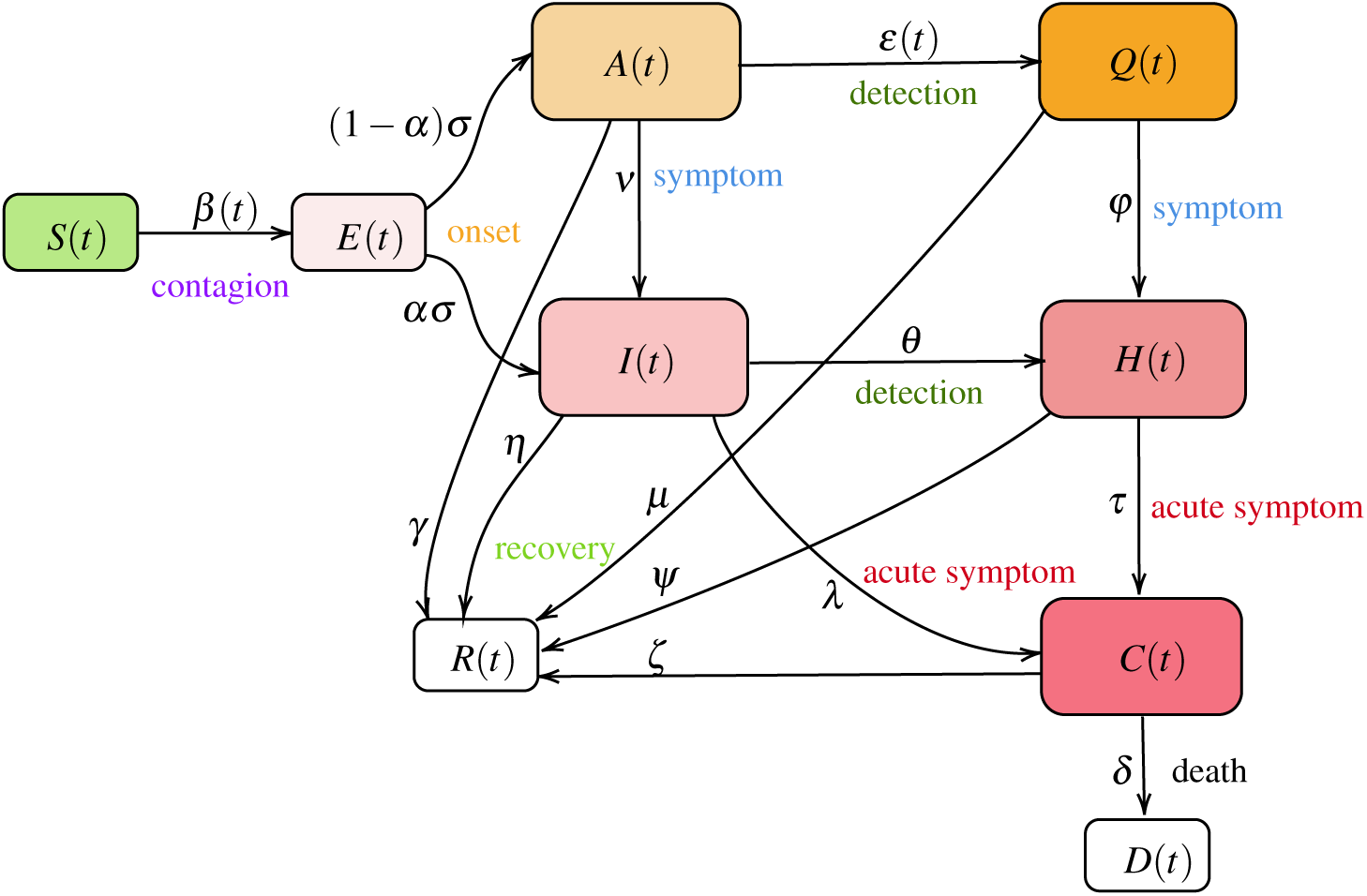
The model consists of following compartments: susceptible *S*(*t*)„ exposed *E*(*t*), asymptomatic *A*(*t*), symptomatic *I*(*t*), quarantined *Q*(*t*), isolated *H*(*t*), deceased (*D*(*t*) and recovered *R*(*t*) individuals in a population of *N*(*t*) = *S*(*t*) + *E*(*t*) + *A*(*t*) + *I*(*t*) + *Q*(*t*) + *H*(*t*) + *R*(*t*) + *D*(*t*) individuals.

The transmission dynamics of COVID-19 in the basic model is given by the following deterministic system of nonlinear differential equations (1)-(10):

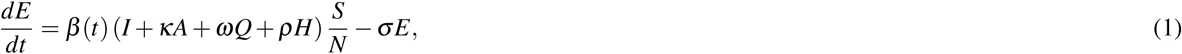

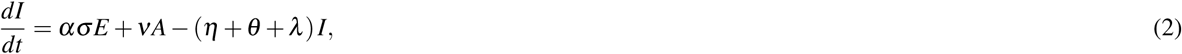

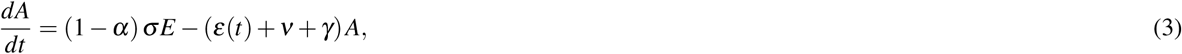

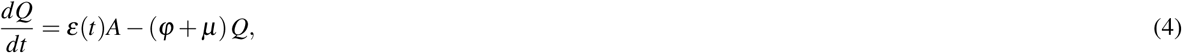

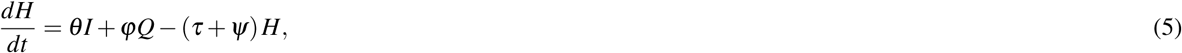

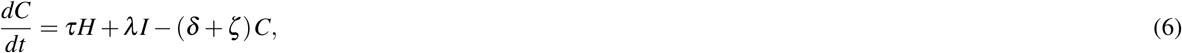

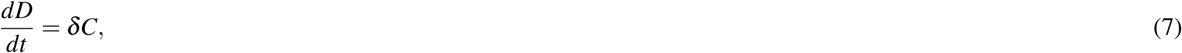

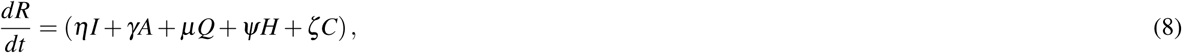

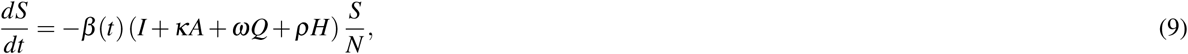

where,

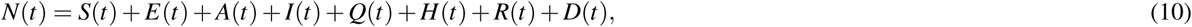

is the total population.

### Susceptible individuals: S(t)

In our model, the susceptible individuals gets exposed to infection, and move to exposed group *E*(*t*), from coming in contact with an infected individual, who may be symptomatic, asymptomatic, quarantined, or isolated. *β* (*t*) is the baseline infectious contact rate, which can vary with time or assumed constant for the analysis of our baseline model. We assume that a person who is infected with symptom, and is not isolated, has the basic transmission coefficient of *β* (*t*), that is changing over time. Based on^11,12^, we define *β* (*t*) to have a value *β*_0_ till time *t*_0_ and then as a decreasing function with respect to time *t*, to reaching *β*_min_.

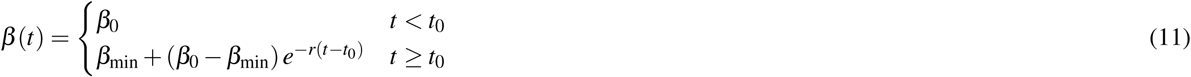

We would like to note that in certain countries, stringency measures were in place at an earlier stages of the epidemic and was relaxed over time leading to a higher contact rate. Under such scenario, we use an increasing function for *β* (*t*) after an initial phase of reaching or nearing *β*_min_ as

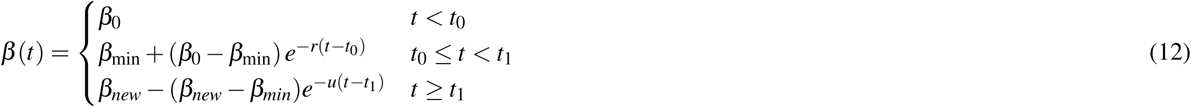

We assume that the asymptomatic individuals infect with a lower contact rate (*κ* < 1) than the symptomatic individuals. Once someone symptomatic is diagnosed, they can only infect healthcare workers and this lower contact rate is captured by the parameter (*ρ* < 1). Similarly, quarantined individuals have much lower contact rate of (*ω* < 1). Overall rate of change for the susceptible population is thus defined by equation (9).

### Exposed individuals: E(t)

Individuals in compartment *E*, are exposed to the virus, and are not contagious during a period of latent time. An individual in *E* becomes infectious, and moves to compartment *A* as asymptomatic or to *I* as symptomatic. We assume that *σ* is the transition rate from exposed to infectious, and a fraction *α* of them show symptoms. Overall rate of change for the exposed population is thus defined by equation (1).

### Symptomatic individuals: I(t)

Symptomatic individuals can get diagnosed (*θ*) and be isolated, or show acute symptoms and be hospitalized (*λ*), or can recover at the rate *η*. It has been observed that *η ≥γ*, where symptomatic individuals recover at faster rate than asymptomatic individuals, and asymptomatic individuals have longer duration of viral shedding^6^. Overall rate of change is given by equation (2).

### Asymptomatic individuals: A(t)

Asymptomatic individuals can eventually show symptoms and move to *I* at rate *ν* or can have a positive diagnosis and move to quarantine. We model testing of asymptomatic population as a function of time as the community testing process ramps up. Testing rate has been captured as *ε*(*t*). Finally, they can recover at the rate *γ*. Overall rate of change is given by equation (3). A note on testing rate is in order. We model it as following:

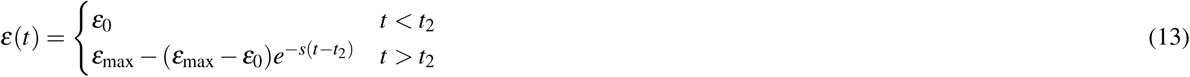

Here, we set the testing rate as an increasing function of time, because of the increasing production of detection kits and the improvement of detection techniques. Where, 1*/ε*_0_ can be thought of waiting time for an individual to get tested in the start of an outbreak and 1*/ε*_max_ is the reduced waiting time at a later point during the epidemic. In other words, lim_*t*→∞_ *ε*(*t*) = *ε*_max_ ≥ *ε*_0_.

### Quarantined individuals: Q(t)

Quarantined individuals are asymptomatic population after diagnosis and they have very low contact rate (*ω* < 1). This is mostly by infecting the other family members or by breach of protocol. They however, may develop symptoms and move to *H* or recover. Overall rate of change is given by equation (4).

### Isolated individuals: H(t)

Isolated individuals are showing symptoms and has been either home isolated or has been hospitalized. They can pass the infection to a limited number of health care professional or caregiver (*ρ*). They can become critical and require treatments in intensive care (*τ*), and a large number of them recover (*ψ*). Overall rate of change is given by equation (5).

### Critical, Recovered and Deceased individuals: C(t), R(t), D(t)

These counters collect information on population that are critical, recovered or have deceased. Overall rate of change is given by equations (6) - (8). We assume that in the base model, recovered individuals possess lasting immunity against SARS-CoV-2 over the period of simulation, however, we extend the model to consider the possibility of re-infection in later section of this paper.

## Results

### Italy

For the COVID-19 epidemic in Italy, we estimate the model parameters based on data from 24th February 2020 (day 1) to 27th August 2020 (day 186) and show the effects of social confinement that is in effect since 9 March 2020, in controlling the spread of the epidemic, and subsequently easing the restriction from 4th May 2020 has again increased contact rate leading to re-emergence. We also model possible longer-term scenarios illustrating the effects of different countermeasures, including social distancing, population-wide testing, vaccination to contain SARS-CoV-2, as well as possible effects of re-infection and loss of immunity.

Model simulation compared to real data is shown in Figure 2. We have estimated *R*_0_ according to equation (23). In our estimate *R*_0_ = 4.09 in the beginning and then reduces to 0.779 after 45 days, and to 0.18 after 90 days. Subsequently, after 110 days it starts increasing and reaches a value 1.67 on day 186.

**Figure 2.**
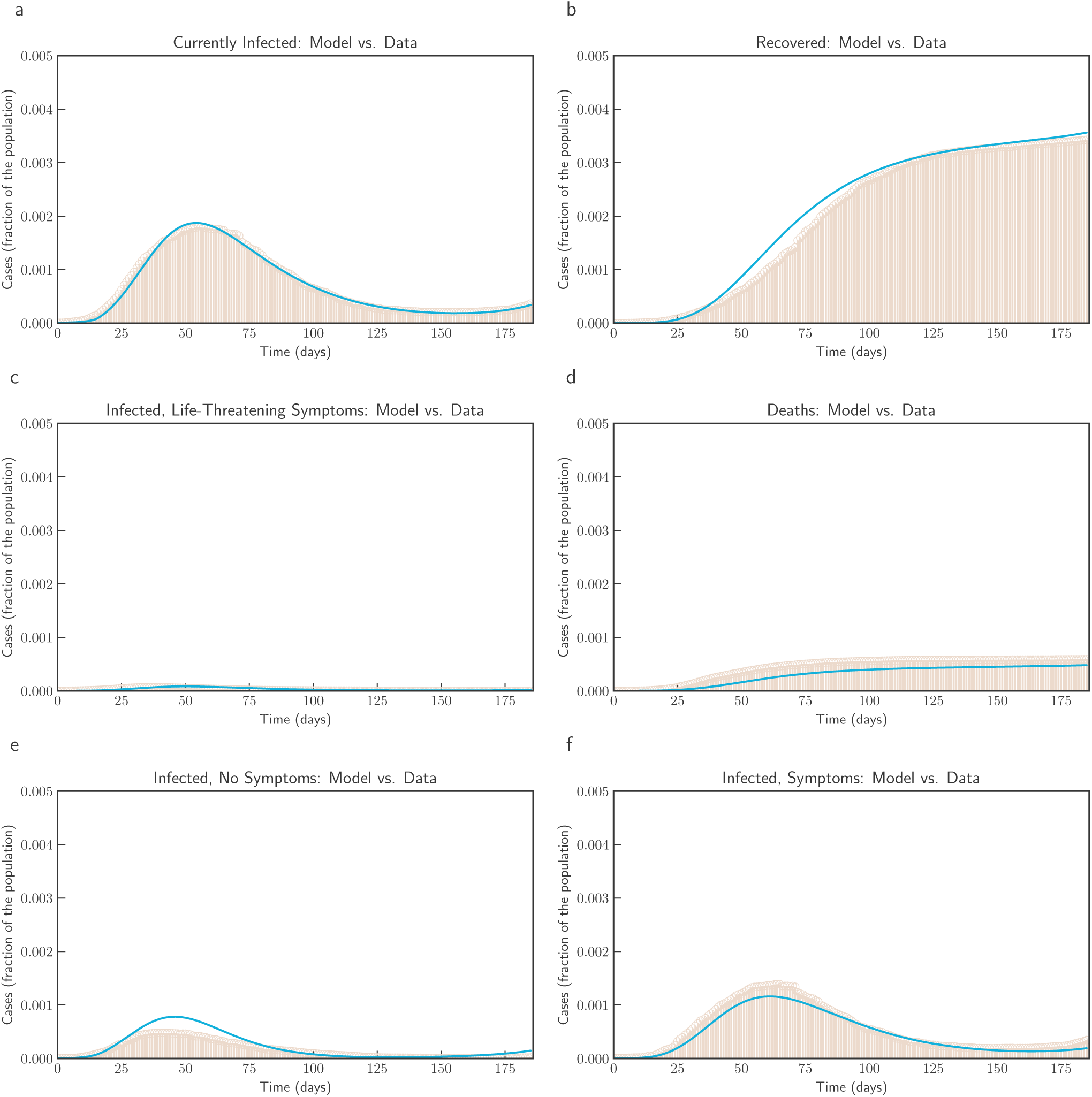
Model simulation compared to real data (Italy) - Comparison between the official data (histogram) and the results with our model. Description of panels: **(a):** Number of currently active cases, (*Q*(*t*) + *H*(*t*) + *C*(*t*)), **(b):** number of reported recovered individuals. 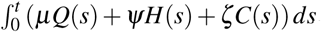, **(c):** number of reported infected with life-threatening symptoms, admitted to ICU, *C*(*t*), **(d):** Number of deceased individuals *D*(*t*), **(e):** number of reported infected with no (or mild) symptoms, who are quarantined at home. *Q*(*t*), **(f):** number of reported infected with symptoms, who are hospitalized. *H*(*t*)

In Figure 3, we show both short term and long term evolution after fitting the model to data. Result shows that without any control measure in long term a significant portion of population getting infected. Hence, we consider the effect of continued social distancing, awareness and confinement measures by sensitivity analysis of contact rate parameter *β* (*t*). We consider reducing *β* (*t*) by 50%, 40%, 30%, 20% and 10% from current contact rate. Figure 4, indicates that reducing contact rate by half can reduce a potential peak infection of 15% population to less than 1.5% of population, and potential deaths from 0.48% to 0.04%.

**Figure 3.**
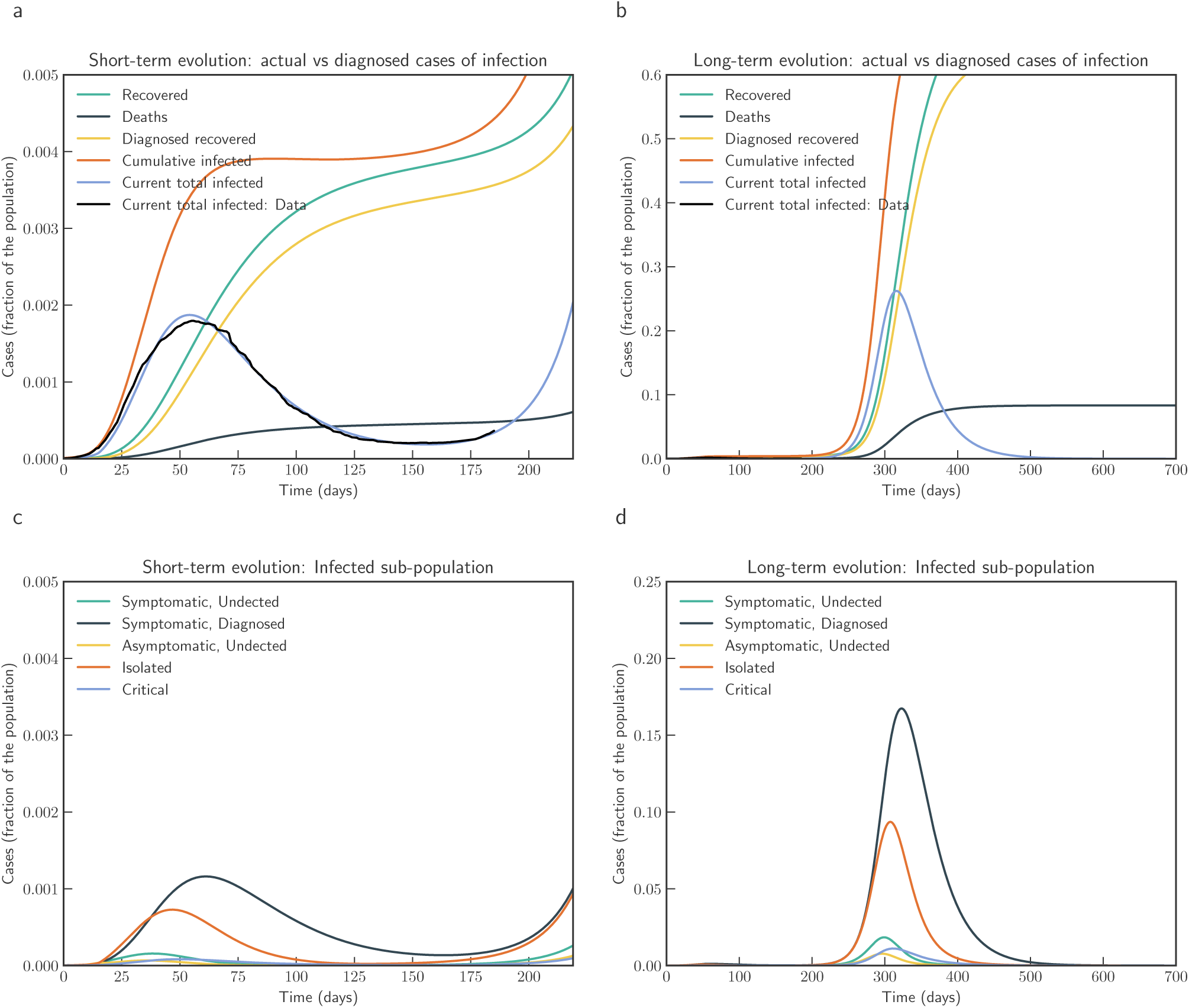
Model simulation compared to real data (Italy) - Epidemic evolution predicted by the model based on the available data. Description of panels: **(a, c):** The short-term epidemic evolution obtained by reproducing the data trend with the model, **(b, d):** Long term epidemic evolution over 700 days. Plots refers to all cases of infection, both diagnosed and non-diagnosed, predicted by the model, although non-diagnosed cases are of course not counted in the data. Note that not all panels are in the same scale.

**Figure 4.**
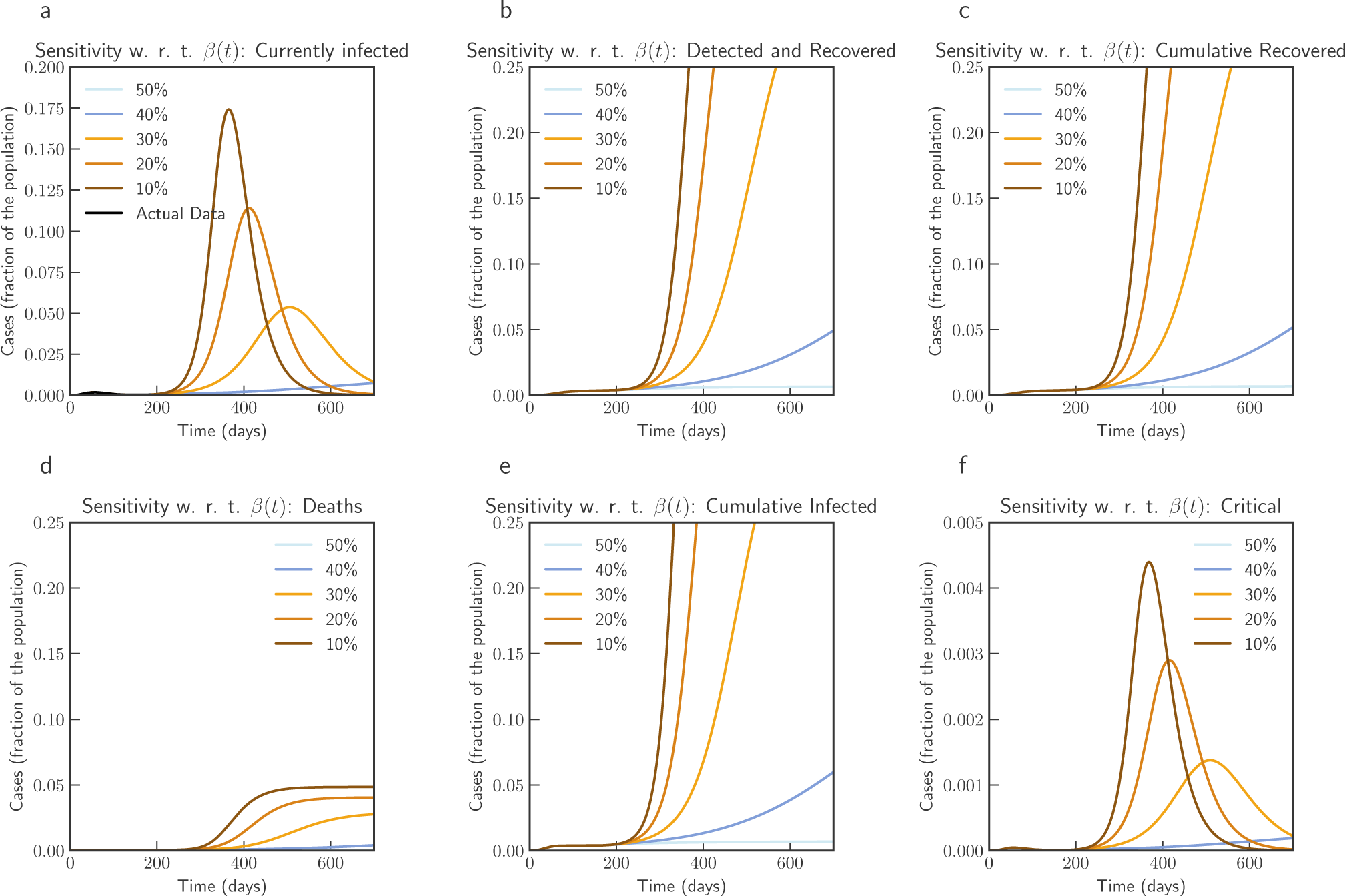
Sensitivity of *β* (*t*) with 50%, 40%, 30%, 20% and 10% reduction from current contact rate (Italy). Note that not all panels are in the same scale.

Subsequently, we also consider the effects of testing and rate of detection of latent cases *ε*(*t*). We consider changing the current testing rate by 2, 4, 6, 8 and 10 times the current rate. Figure 5, the simulation of the model after fitting current data indicates that an increased testing rate by 10 times will reduce potential peak infection rate from 18% of population to 14%, deaths from 0.48% to 0.42%. Our model confirms that extensive testing campaigns can reduce the infection peak (as the diagnosed population enters quarantine and is therefore less likely to affect the susceptible population) and help end the epidemic more quickly. However, it can be observed that sensitivity of contact rate is much more significant than testing. This is also due to that fact that in our model we have considered *ε* as a parameter that impacts how asymptomatic cases are moved to quarantine, while infection from asymptomatic cases are lower compared to symptomatic cases.

**Figure 5.**
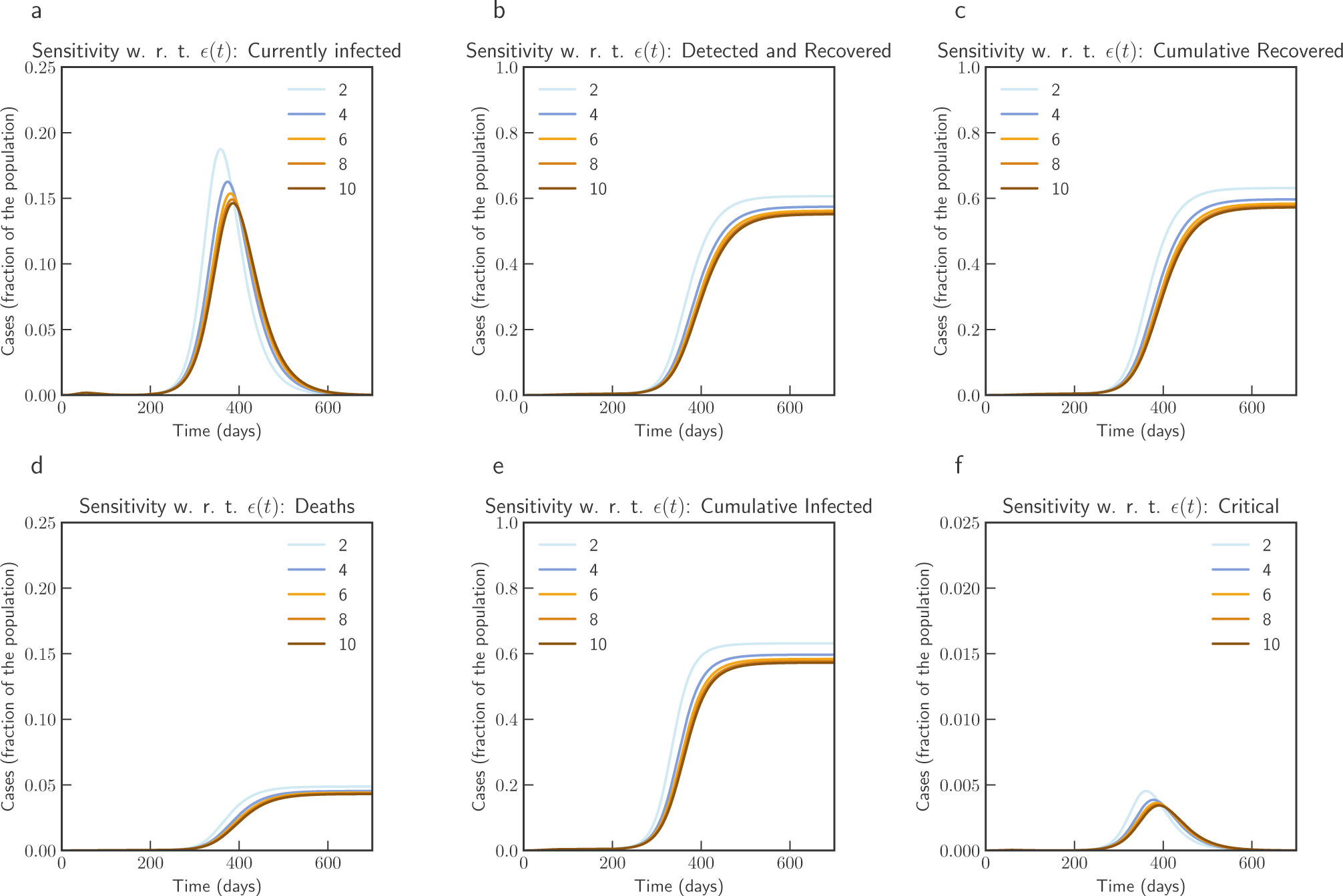
Sensitivity of *ε* (t) with changed rate of testing by 2;4;6;8 and 10 times the current rate (Italy). Note that not all panels are in the same scale.

**Figure 6.**
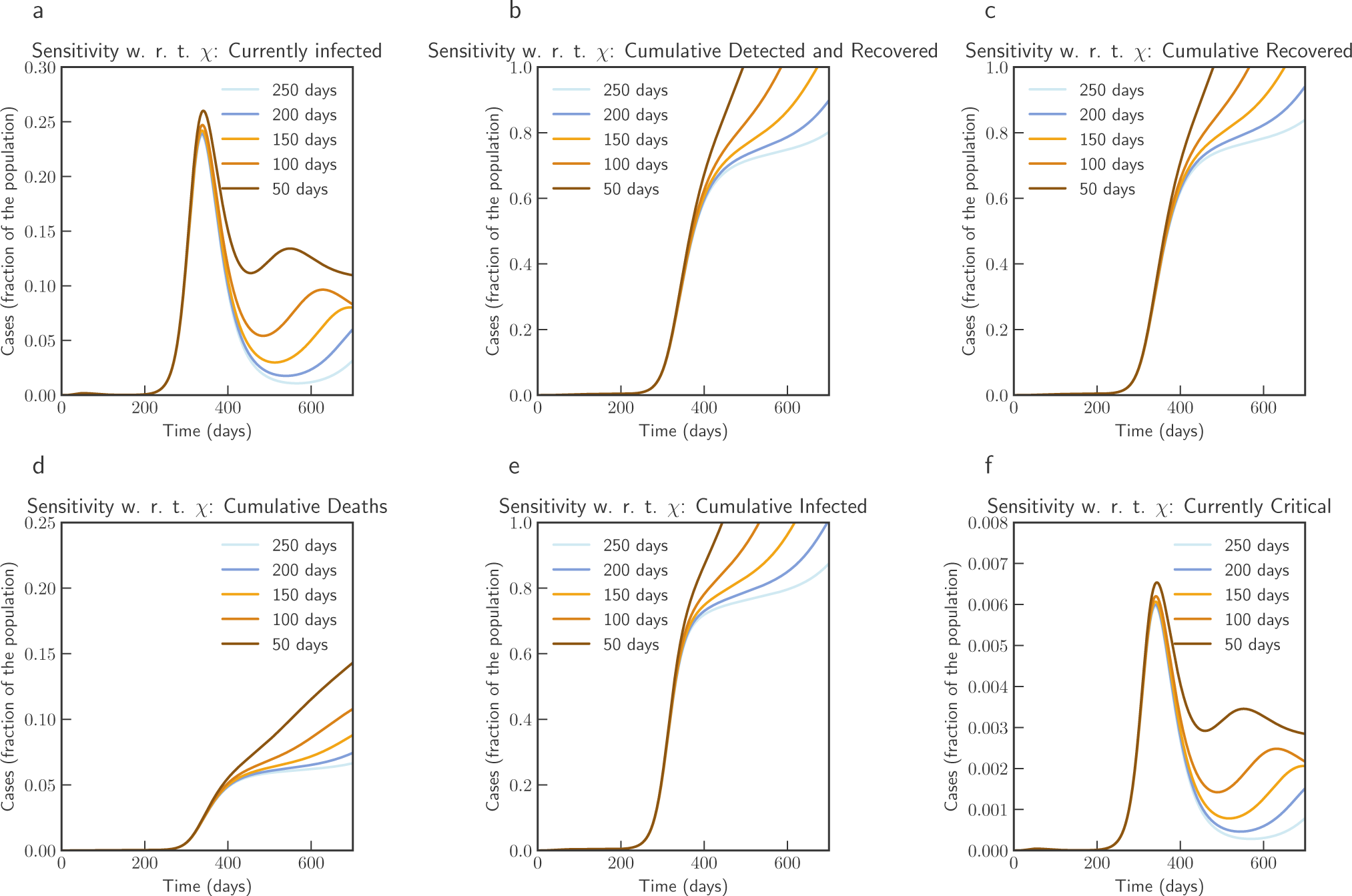
Sensitivity of *χ*(*t*) with loss of acquired immunity over time by 250, 200, 150, 100 and 50 days (Italy). Note that not all panels are in the same scale.

As described by extended model in equation - (32) and (33), we consider the possibility of losing acquired immunity over time and having a reinfection, we use parameter *χ*(*t*) to capture this. In our simulation we consider that *χ* varies from 1*/*250 to 1*/*50. This can be thought as number of days to lose immunity and hence corresponds to 250 days to 50 days. As expected, the simulation shows that if loss of immunity occurs within 3 months, there is a significant chance of subsequent waves of epidemic.

Finally, we simulate mass vaccination scenarios (see Figure 7 and Figure 8) according to equation (35), (36), (37), (38) and (34). We assume that vaccine is available after 90 days from 27th August 2020. *ϕ* is varied from 1.0 (No vaccination), 0.5 (50% efficient), 0.25 (75% efficient), 0.125(87.5% efficient), and 0.0625 (93.75% efficient) and *ξ*_max_ = 0.5 and *ξ*_max_ = 0.9. It can be concluded that even a 75% efficient vaccine can be significantly effective in reducing the impact.

**Figure 7.**
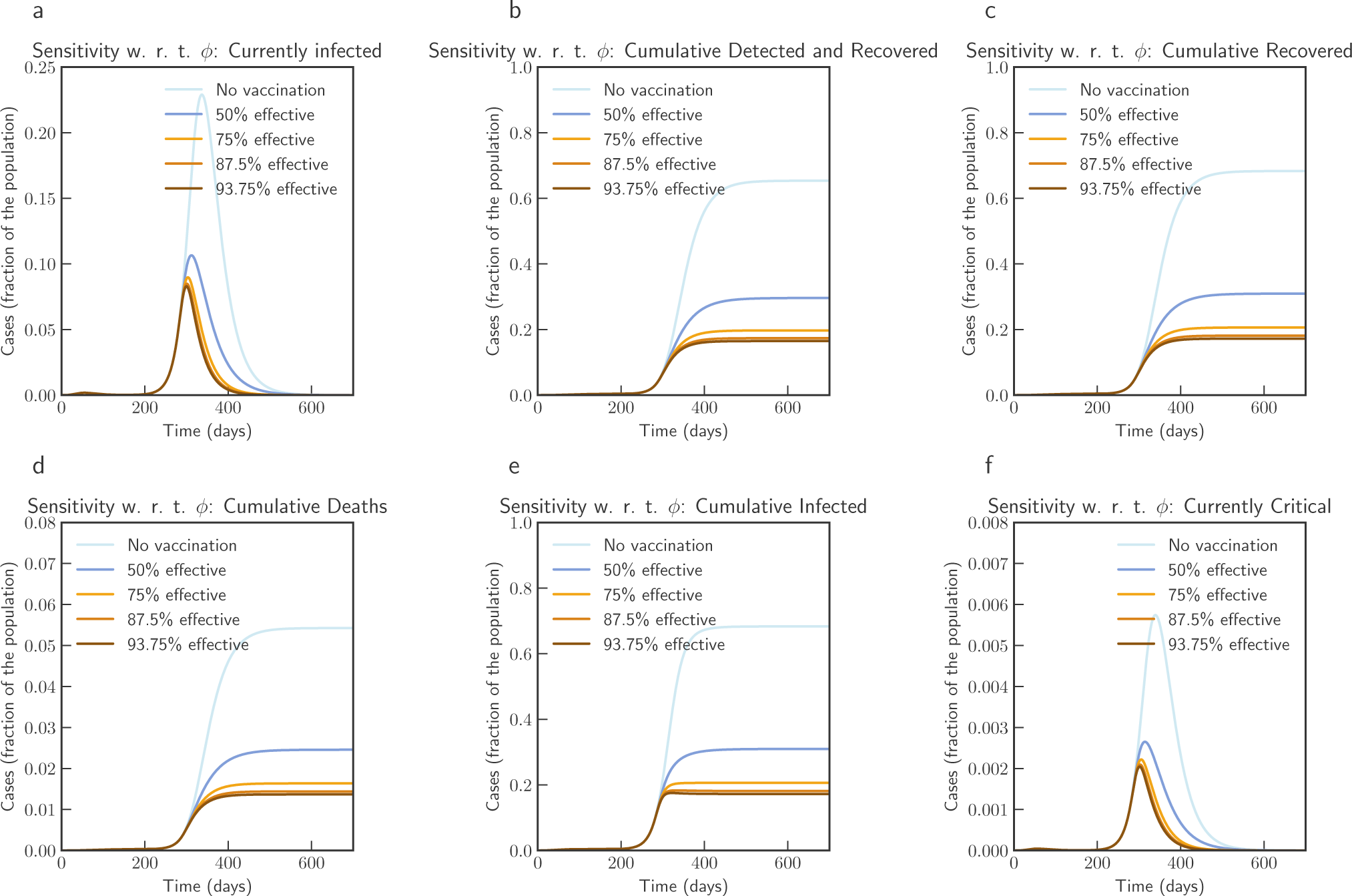
Sensitivity of vaccine efficiency parameter (1 − *ϕ*), where *ϕ*= 0 represents vaccine that offers 100% protection against infection, and *ξ*_max_ = 0.5, i.e. maximum of 50% population is administered with vaccine. *ϕ* is varied from 1.0 (No vaccination), 0.5 (50% efficient), 0.25 (75% efficient), 0.125(87.5% efficient), and 0.0625 (93.75% efficient) (Italy)

**Figure 8.**
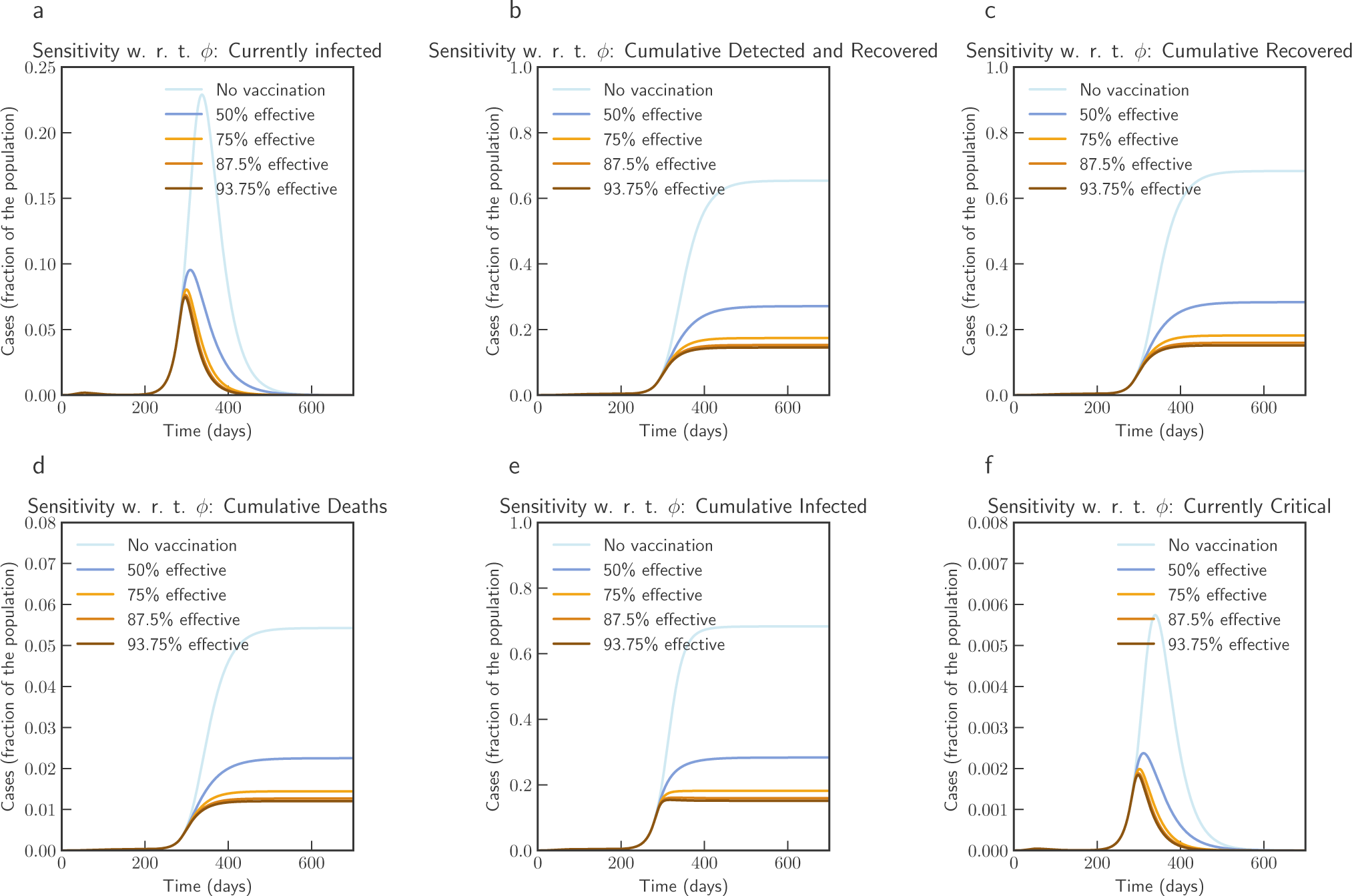
Sensitivity of vaccine efficiency parameter (1 − *ϕ*), where *ϕ*= 0 represents vaccine that offers 100% protection against infection, and *ξ*_max_ = 0.9. *ϕ* is varied from 1.0 (No vaccination), 0.5 (50% efficient), 0.25 (75% efficient), 0.125(87.5% efficient), and 0.0625 (93.75% efficient) (Italy)

### India

India had one of the most strict stay home order across the country in first phase of the lock-down between 25 March 2020 – 14 April 2020 (21 days), where an entire population of 1.3 billion people was put under restricted movement. Overall the lock-down had multiple phases, second phase was from 15th of April 2020 to 3rd of May 2020, and third phase was 4th of May to 17th of May, 2020.

We use data from Johns Hopkins University Center for Systems Science and Engineering (JHU CSSE). We use data from 2nd March 2020 (Day 1) to 16th September 2020 (Day 198). Figure 9 compares the fitted model with actual data. Based on our estimate, initial value of *R*_0_ was 1.762, which reduces to 1.68 after a month when lockdown as in place. At the end of lockdown phases it is about 1.5, and finally after 194 days it reaches 1.03. These values are comparable with findings in^13,14^.

**Figure 9.**
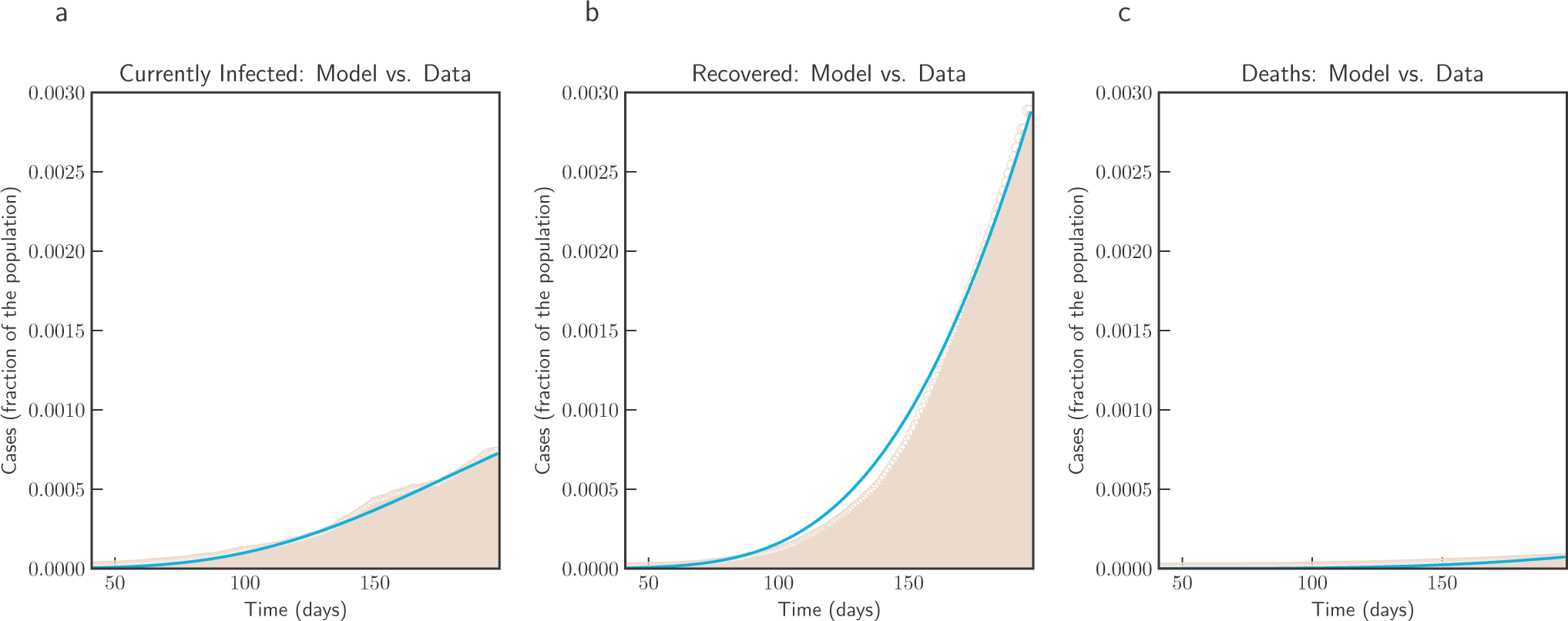
Model simulation compared to real data (India) - Comparison between the official data (histogram) and the results with our model. Description of panels: **(a):** Number of currently active cases, (*Q*(*t*) + *H*(*t*) + *C*(*t*)), **(b):** number of reported recovered individuals. 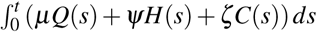, **(c):** Number of deceased individuals *D*(*t*)

We have simulated the long term evolution of the epidemic for India after estimating the parameters as shown in Figure 10. Simulation suggests that peak in India will be reached around 275 days from 2nd March 2020 with about 2.6% population getting infected.

**Figure 10.**
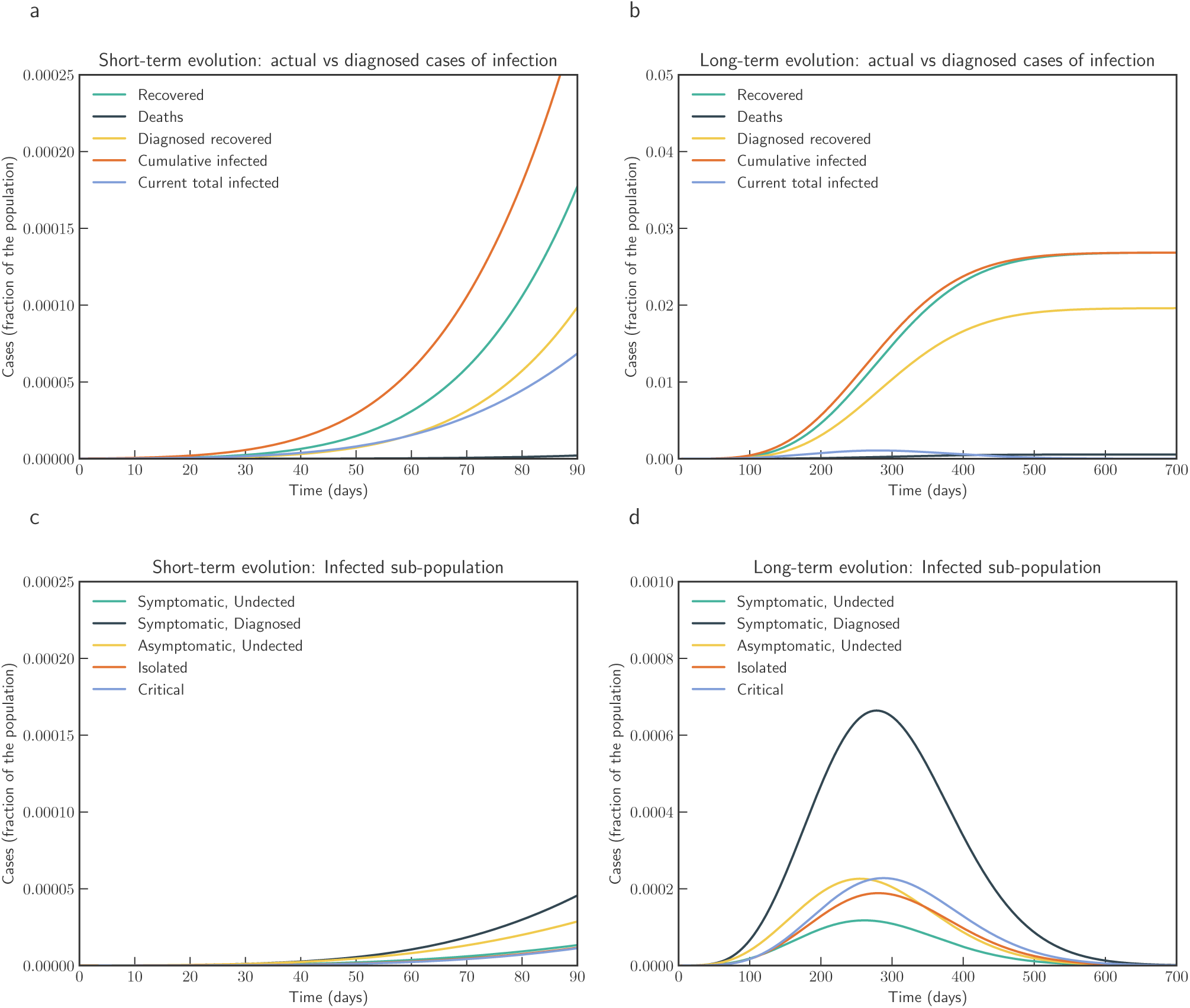
Model simulation compared to real data (India) - Epidemic evolution predicted by the model based on the available data. Description of panels: **(a, c):** The short-term epidemic evolution obtained by reproducing the data trend with the model for 90 days, **(b, d):** Long term epidemic evolution over 700 days. Plots refers to all cases of infection, both diagnosed and non-diagnosed, predicted by the model, although non-diagnosed cases are of course not counted in the data. Note that not all panels are in the same scale.

Simulation studies on impact of reducing contact rate and testing is captured in Figure 11 and Figure 12 respectively. From the simulation we can observe that the sensitivity to contact rate is higher compared to increased testing. A reduction in contact rate by 50% compared to a reduction of 10% can reduce death from 0.0268% to 0.0141% of population.

**Figure 11.**
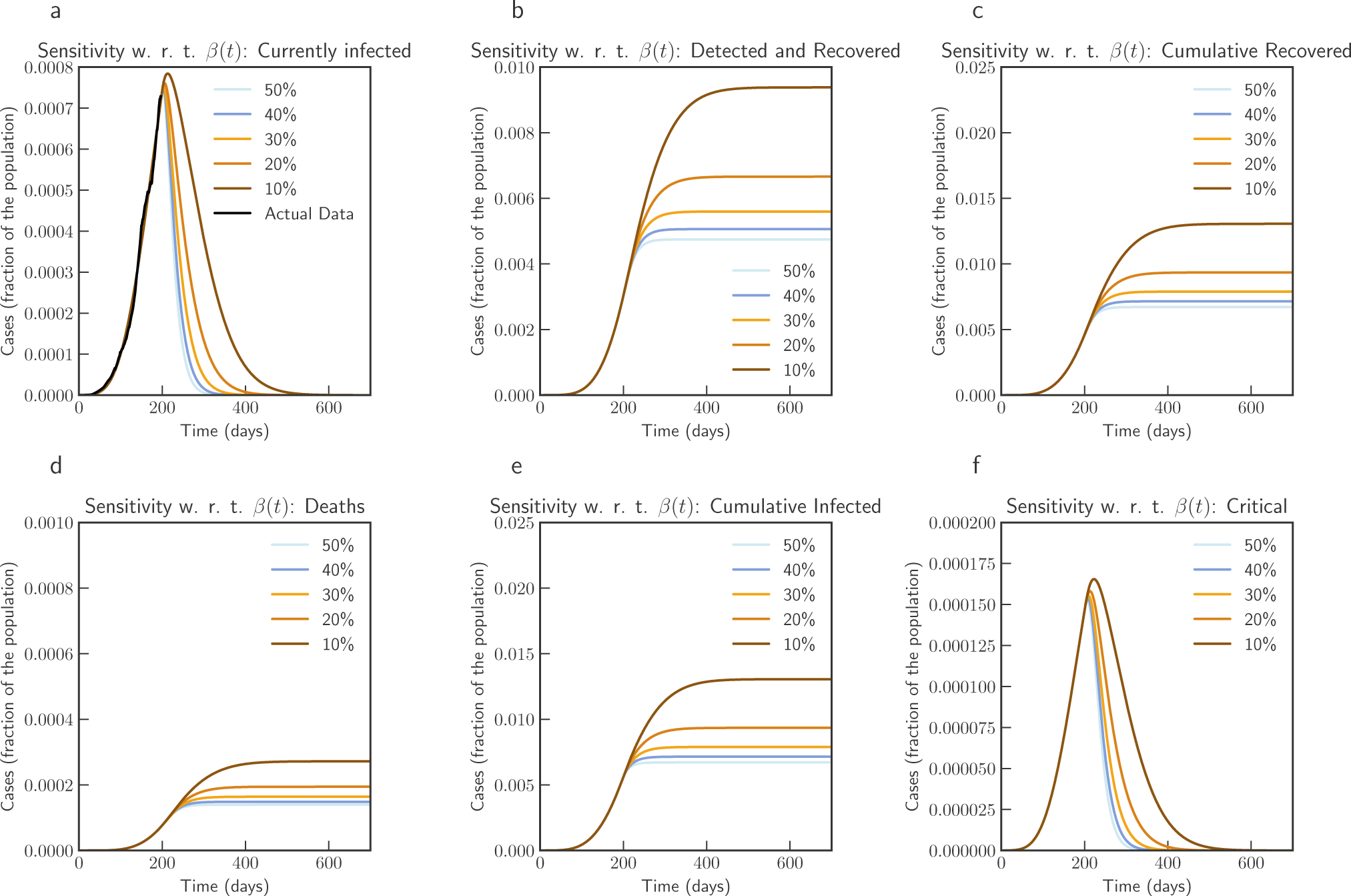
Sensitivity of *β* (*t*) with 50%, 40%, 30%, 20% and 10% reduction from current contact rate (India). Note that not all panels are in the same scale.

**Figure 12.**
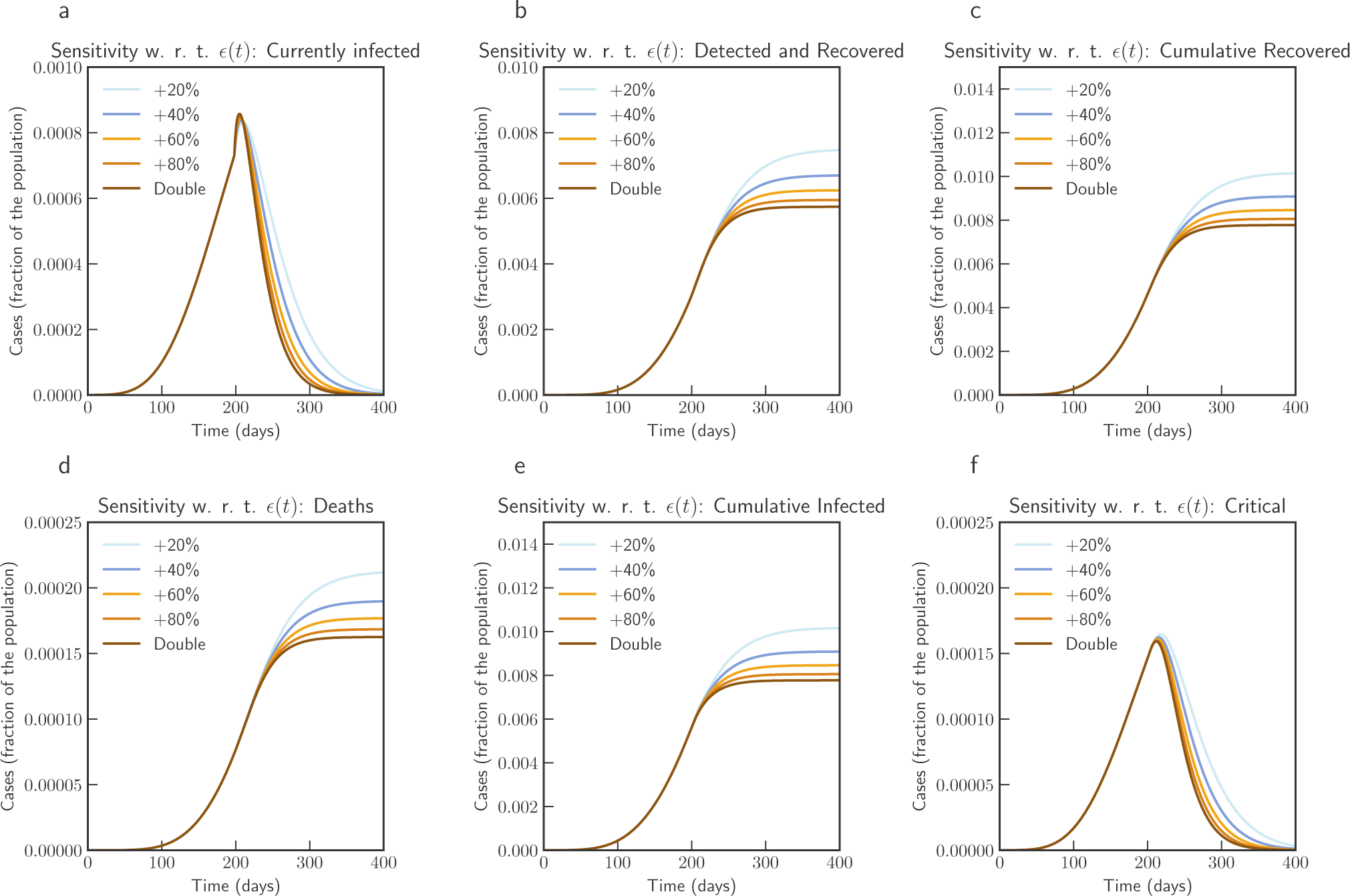
Sensitivity of *ε*(*t*) with changed rate of testing by 1.2, 1.4, 1.6, 1.8 and 2 times the current rate (India). Note that not all panels are in the same scale.

Figure 13 show the simulation study loss of acquired immunity for scenario in India. We note that this model is not very sensitive to *χ*(*t*). This is due to the size of the population and current state of the epidemic in India - reinfection within 2 months changes total number of infected population from 2.66% to 2.98%, while reinfection after 250 days changes this to 2.768%.

**Figure 13.**
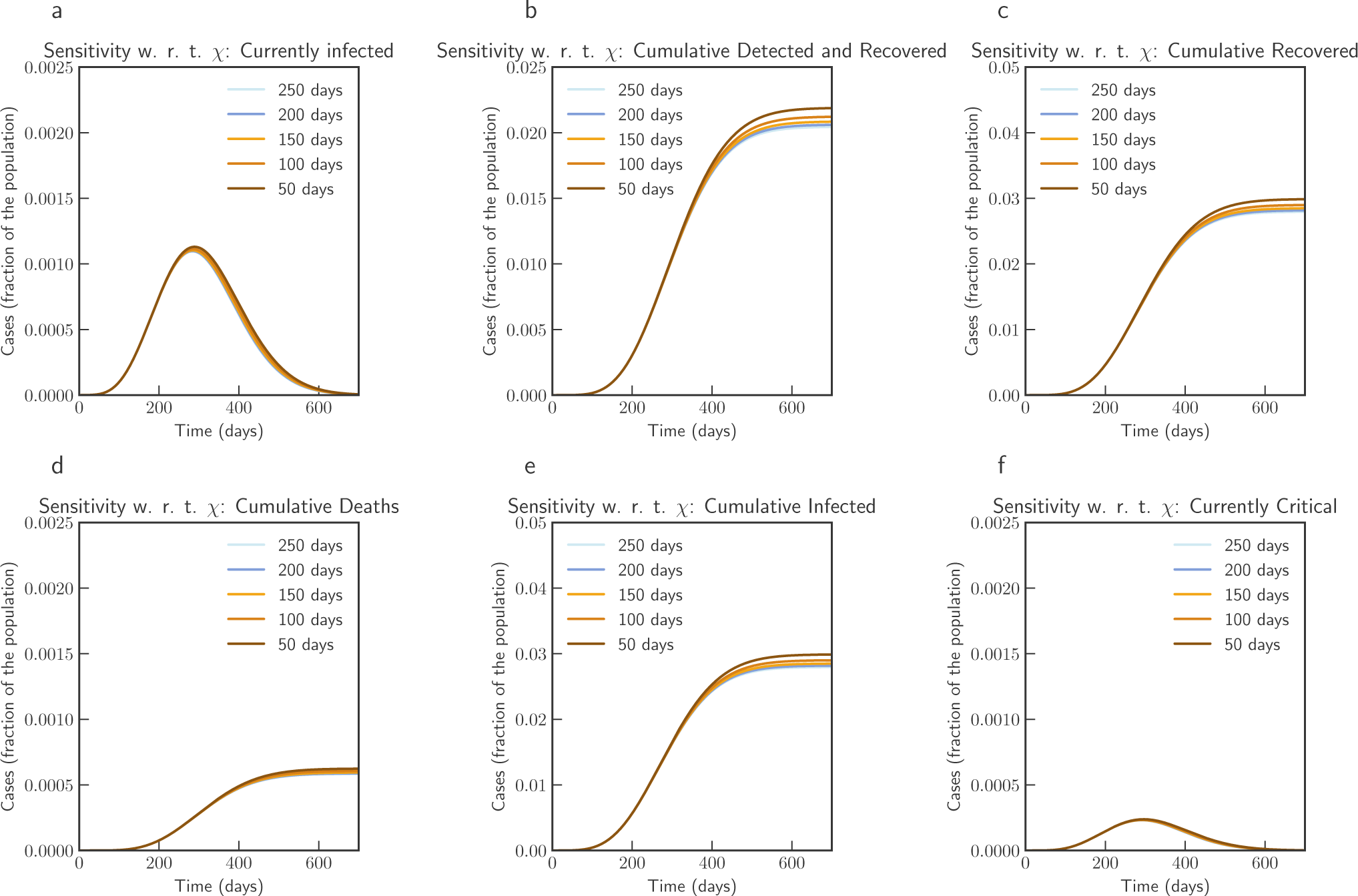
Sensitivity of *χ*(*t*) with loss of acquired immunity over time by 250, 200, 150, 100 and 50 days (India). Note that not all panels are in the same scale.

Simulation studies for vaccination is presented in Figure 14 and Figure 15. We assume that the vaccine is available from 90 days after 16th September 2020. Simulation suggests that a 0.056% of population would die without vaccination, while 93.75% efficient vaccine given to 30% population would bring this down to 0.036% of population, and 93.75% efficient vaccine given to 70% population would bring this down to 0.034%.

**Figure 14.**
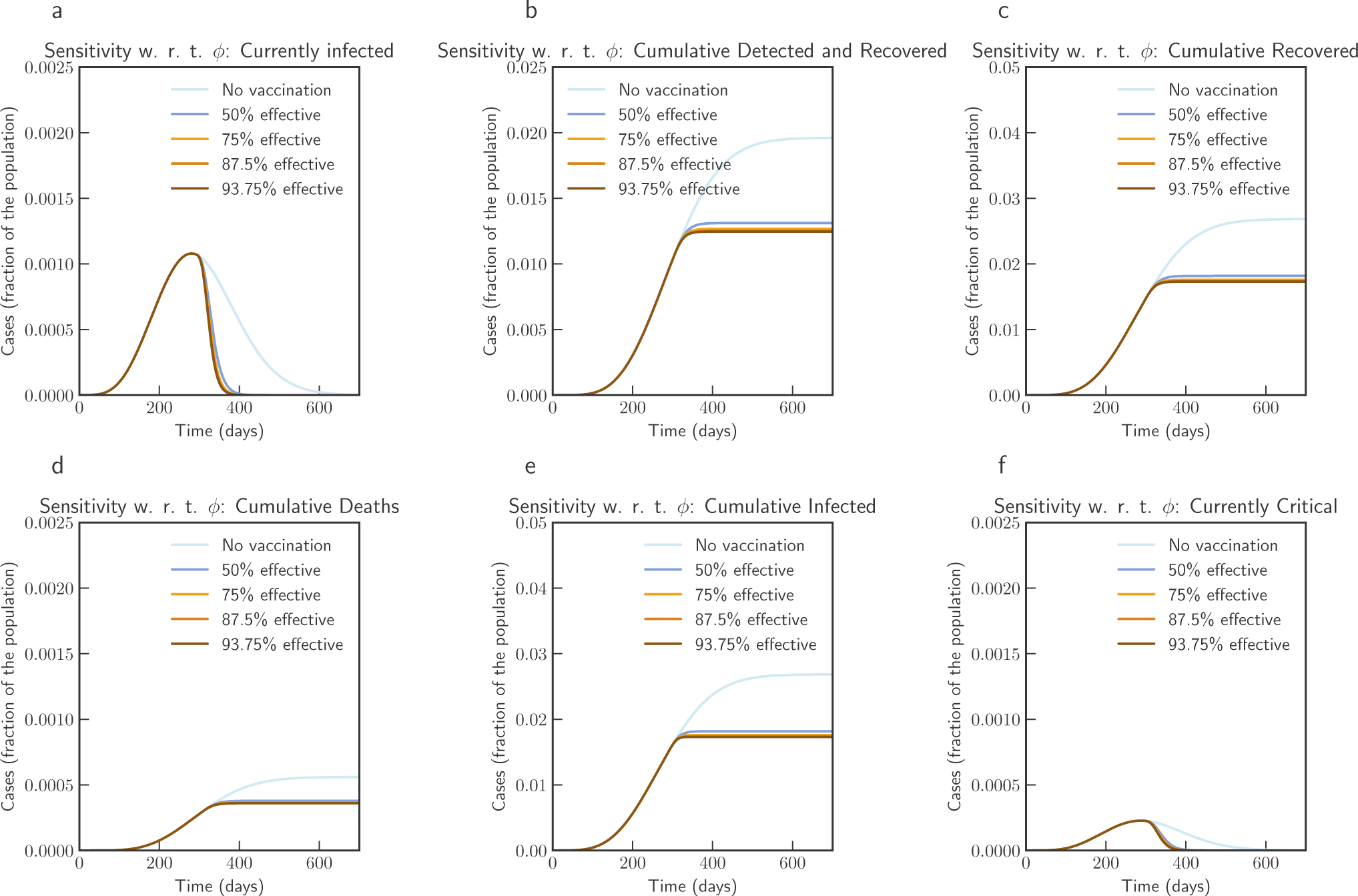
Sensitivity of vaccine efficiency parameter (1 − *ϕ*), where *ϕ* = 0 represents vaccine that offers 100% protection against infection, and *ξ*_max_ = 0.3, i.e. maximum of 30% population is administered with vaccine. *ϕ* is varied from 1.0 (No vaccination), 0.5 (50% efficient), 0.25 (75% efficient), 0.125(87.5% efficient), and 0.0625 (93.75% efficient) (India)

**Figure 15.**
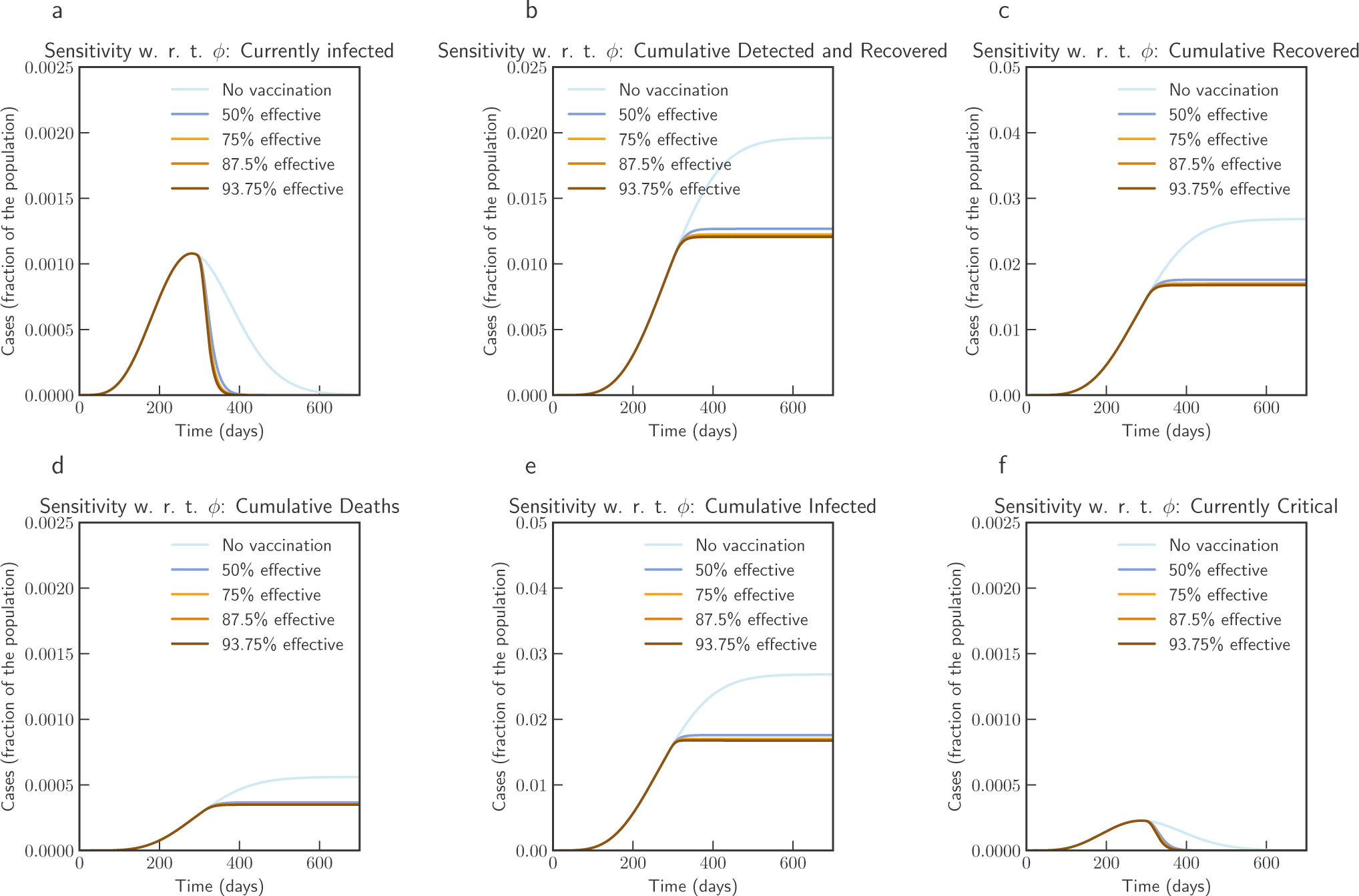
Sensitivity of vaccine efficiency parameter (1 − *ϕ*), where *ϕ* = 0 represents vaccine that offers 100% protection against infection, and *ξ*_max_ = 0.7, i.e. maximum of 70% population is administered with vaccine. *ϕ* is varied from 1.0 (No vaccination), 0.5 (50% efficient), 0.25 (75% efficient), 0.125(87.5% efficient), and 0.0625 (93.75% efficient) (India)

### Victoria, Australia

For Victoria, Australia we use data from 27 February, 2020 (day 1) to 9 September, 2020 (day 195). Consistent with other places Victoria had initial outbreak during March and April 2020 - which was effectively controlled. Control measures were relaxed during June 2020 to revive economic activities. Based on our estimate *R*_0_ in Victoria was 2.72 in the beginning of July. A stricter confinement was placed in selected localities on 1 July 2020, which was extended to the whole of metropolitan Melbourne and Mitchell Shire on 8 July. Estimated by our model, *R*_0_ was 2.86 at that time. However, the restriction was effective to bring down *R*_0_ to 1.13 by 30, July, 2020. We note that our findings are similar to^15^. In our estimate, current *R*_0_ is well below 1.

## Methods

### Baseline mathematical models

#### Baseline epidemiological parameters

In this section we describe the estimated values of various parameters based on the current literature. It has been noted in the literature that the clinical course of the disease is typically quite long. Average total duration of illness has been estimated to be three weeks in^16^. Parameter *β*_0_ is strongly dependent on the population behavior. We select a default value that has been estimated in^17^ for pre-lockdown period. Baseline values for various parameters are listed in the Table - 1.

**Table 1.** Baseline parameters, brief description, possible ranges based on modeling and clinical studies, and default value chosen for this study. Note that these parameters were estimated to fit the model to real data

| Param | Description | Possible Range | Default |
| --- | --- | --- | --- |
| $\beta$ | infectious contact rate | 0.5-1.5 $day^{-1}$<sup>17,18</sup> | 1.14 $day^{-1}$ |
| $\kappa$ | infectiousness factor asymptomatic | 0.4-0.6<sup>18,19</sup> | 0.5 |
| $\omega$ | infectiousness factor quarantined | 0.005-0.0114<sup>9</sup> | 0.0114 |
| $\rho$ | infectiousness factor isolated | 0.005-0.0114<sup>9</sup> | 0.0114 |
| $\sigma$ | transition rate exposed to infectious | 1/14-1/3 $day^{-1}$<sup>18,20</sup> | 1/5.2 $day^{-1}$ |
| $\alpha$ | fraction of infections that become symptomatic | 0.15-0.7<sup>18,19,21</sup> | 0.3 |
| $\nu$ | transition rate asymptomatic to symptomatic | 0.025-0.125<sup>9</sup> | 0.125 |
| $\varepsilon$ | detection rate asymptomatic | 0.171<sup>9</sup> | 0.171 |
| $\varphi$ | rate of quarantined to isolation | 0.025-0.125<sup>9</sup> | 0.125 |
| $\theta$ | rate of detection of symptomatic | 0.371<sup>9</sup> | 0.371 |
| $\tau$ | rate of developing life-threatening symptoms in isolation | 0.027<sup>9</sup> | 0.027 |
| $\lambda$ | rate of developing life-threatening symptoms for symptomatic | 0.017<sup>9</sup> | 0.017 |
| $\gamma$ | recovery rate of asymptomatic | 0.034<sup>9</sup> | 0.034 |
| $\eta$ | recovery rate of symptomatic | 0.017<sup>9</sup> | 0.017 |
| $\mu$ | recovery rate of quarantined | 0.034<sup>9</sup> | 0.034 |
| $\psi$ | recovery rate of isolated | 0.017<sup>9</sup> | 0.017 |
| $\zeta$ | recovery rate of critical | 0.017<sup>9,21</sup> | 0.017 |
| $\delta$ | mortality rate | 0.01-0.05<sup>9,19,22</sup> | 0.01 |

#### The basic reproduction number for baseline model

The basic reproduction number is calculated for the special case when we have *β* (*t*) = *β*_0_, *ε*(*t*) = *ε*_0_. In following we explore the local stability of the disease-free equilibrium (DFE) using the next generation operator method^23,24^. Following^24^, we define the system of equations (1)-(10), in more compact form as:

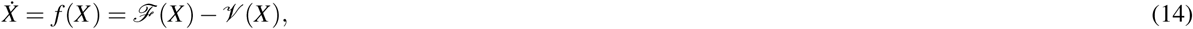

where, *X* = (*E, I, A, Q, H,C, D, R, S*)^*t*^, and ℱ (*X*) containing rate of appearance of new infections defined as:

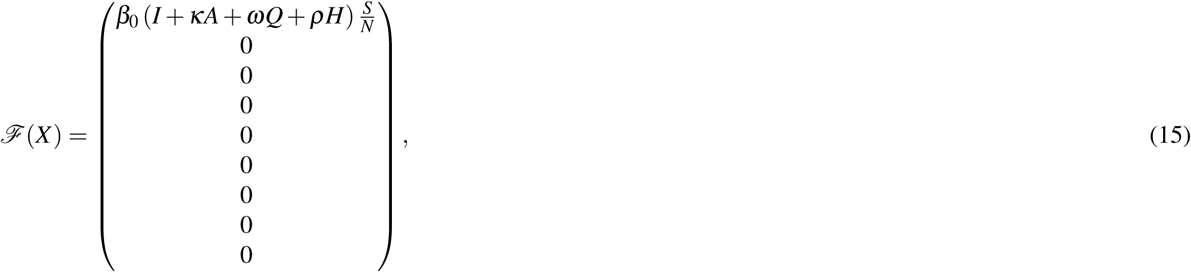

and, 𝒱 (*X*) capturing the movement between the compartments, with 𝒱 ^−^(*X*) as the rate of outward transfer, and 𝒱 ^+^(*X*) as the rate of inward transfer for each compartment, we have,

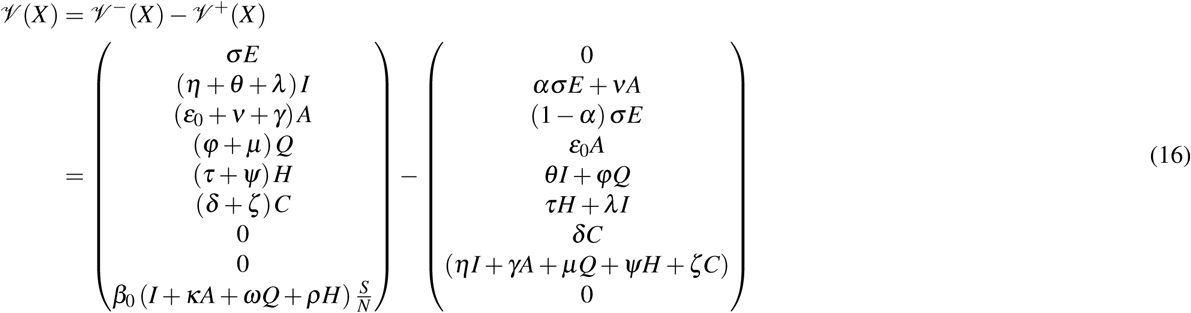

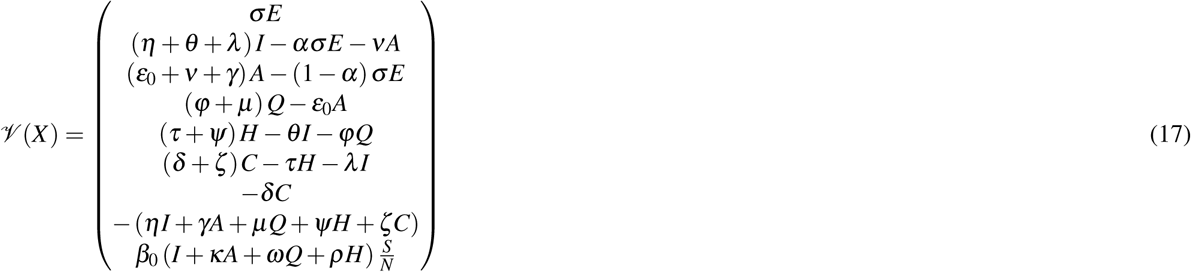

We also define 𝒳_*s*_, as the set of all possible disease free states. In order to directly apply the results in^24^, following shall hold for equation 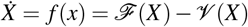:

1. Functions ℱ (*X*), 𝒱 ^−^(*X*) and 𝒱 ^+^(*X*), are all non-negative, when *X* > 0.
2. If *X* ∈ 𝒳_*s*_, then 𝒱 ^−^(*x*) = ℱ (*x*) = 𝒱 ^+^(*x*) = 0 for *x* ∈ {*E, I, A, Q, H,C, D, R*}.
3. Let *D f* (*X*_0_) be the Jacobian matrix evaluated at DFE *X*_0_, and defined as the partial derivative [*∂ f /∂x*] for *x* ∈ {*E, I, A, Q, H,C, D, R, S*}. If ℱ (*X*) = 0, then all eigenvalues of *D f* (*X*_0_) has negative real parts.

We note that each function represents a directed transfer of individuals, and they are all non-negative. (1) and (2), can be observed from the equations (15) and (17). For (3), setting ℱ (*X*) = 0, we consider linearized system 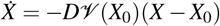, near DFE. From equation (20) we observe that eigenvalues corresponding to *D f* (*X*_0_) has zero eigenvalues of multiplicity 3 with associated eigenvectors in the directions of *D, R, S*. The results in^24^ still holds for our system for stability in the directions of the susceptible and recovered compartment (note that)as *D* is a counting compartment), this however, has no consequence in the meaning of the threshold ℛ_0_. In fact this technicality can be resolved by adding natural birth and death rates proportional to the compartments *S* and *R* that is arbitrarily small and positive. Let *X*_0_ ∈ 𝒳 _*s*_ be a DFE. Then *X*_0_ = (0, 0, 0, 0, 0, 0, 0, 0, *S*_0_), and

with *S*_0_*/N*_0_ = 1 we have,

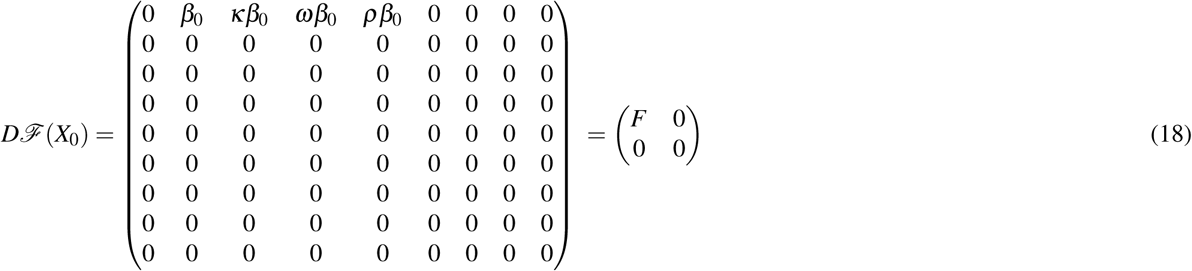

With,

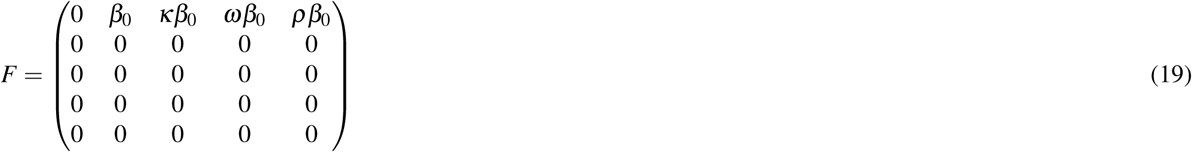

Similarly, we have

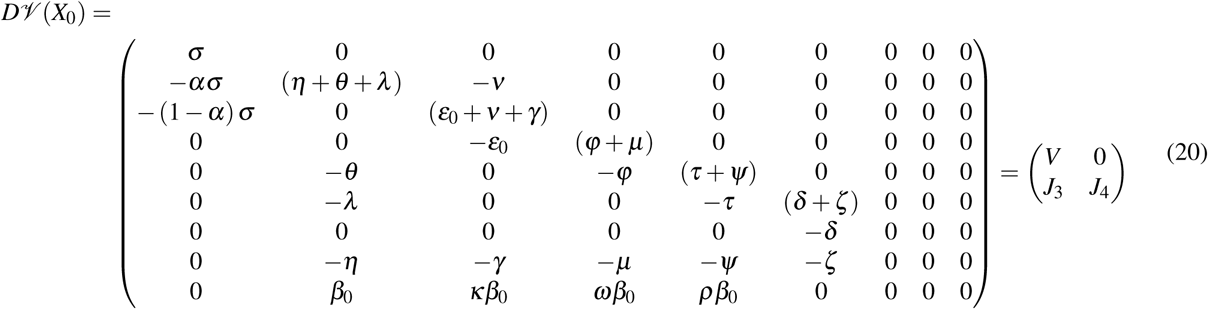

With,

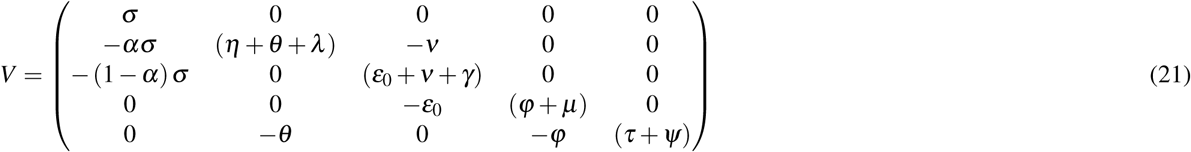

and,

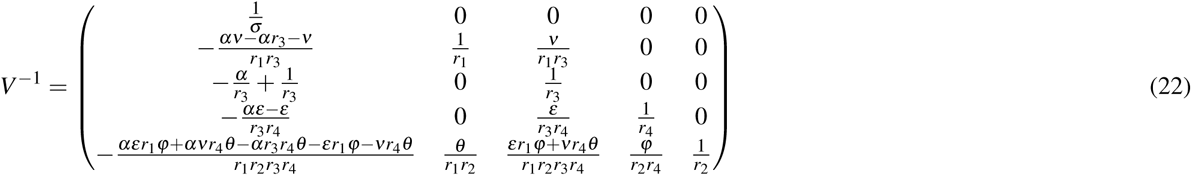

Defining *ρ*(*A*) = max {| *λ*_1_ |, …, |*λ*_*n*_} as the spectral radius of an *n*×*n* matrix *A*, with eigenvalues *λ*_1_ … *λ*_*n*_, and | | denoting absolute values. According to^24^, basic reproduction number ℛ_0_ associated to the system can be computed as ℛ_0_ = *ρ*(*FV* ^−1^). Hence, based on the discussion above, for the baseline model we have,

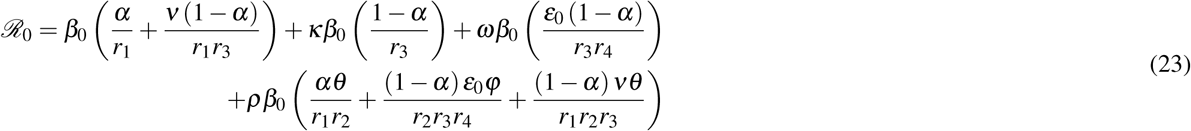

where, *r*_1_ = (*η* + *θ* + *λ*), *r*_2_ = (*τ* + *ψ*), *r*_3_ = (*ε*_0_ + *ν* + *γ*), and *r*_4_ = (*φ* + *µ*). When the epidemic is over, we will have the condition that 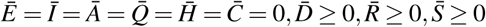, with 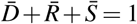. That is, only the susceptible, the recovered and the deceased individuals are eventually present.

##### Proposition 1

*The system of equation with susceptible population* 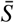 *is asymptotically stable if and only if*

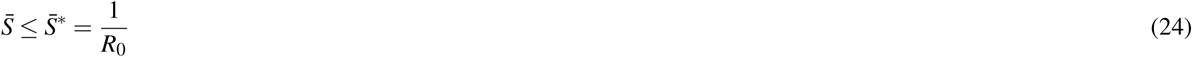

*Proof*. The dynamical system matrix of the linearized system near a DFE is given by:

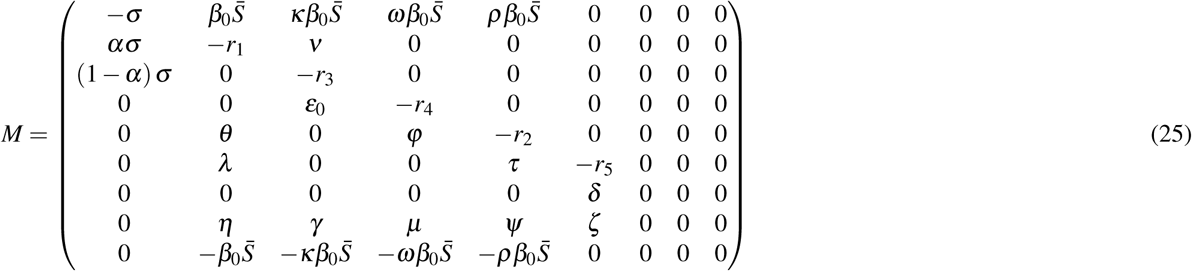

Where *r*_1_ = (*η* + *θ* + *λ*), *r*_2_ = (*τ* + *ψ*), *r*_3_ = (*ε*_0_ + *ν* + *γ*), *r*_4_ = (*φ* + *µ*). and *r*_5_ = (*δ* + *ζ*). Using det(*sI−M*) = 0, we have the characteristic polynomial of *M* having following form: *s*^3^ (*r*_5_ + *s*) *p*(*s*). Hence, the matrix has three null eigenvalues, one eigenvalue of −(*δ* + *ζ*), and five eigenvalues that are roots of the polynomial *p*(*s*), where,

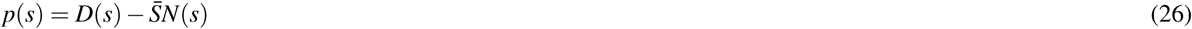

with,

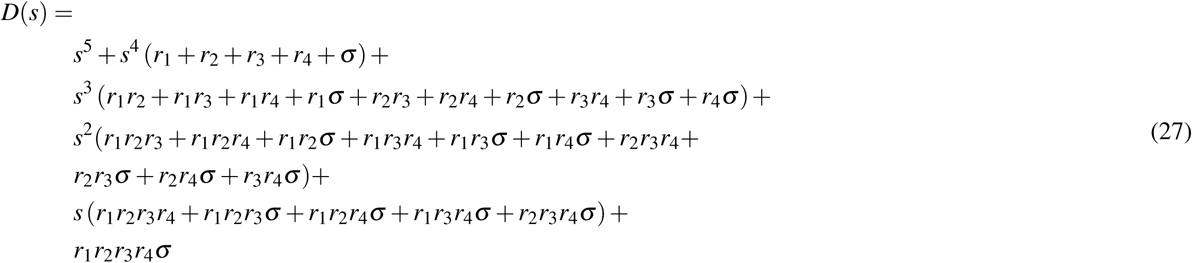

and,

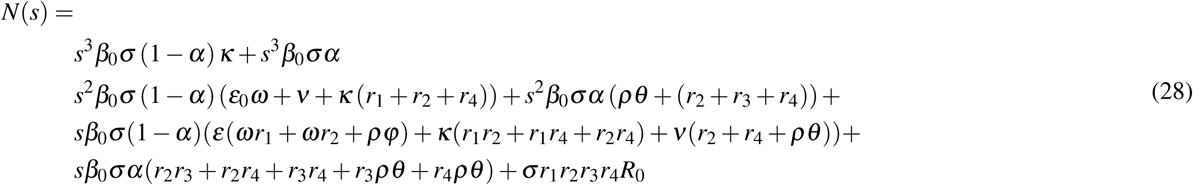

Defining *G*(*s*) = *N*(*s*)*/D*(*s*), and noting that the system is positive, and hence *H*_∞_ norm of *G*(*s*) is equal to the static gain *G*(0) = *N*(0)*/D*(0) = *R*_0_. To have the real part of every root of the polynomial to be zero or negative (Hurwitz), we should have 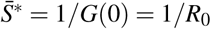. Note that the stability of the equilibrium occurs for 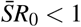. Since in the beginning of an epidemic we have 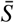 very close to 1, it should be noted that *R*_0_ < 1 is required to have small effect and stability.

### Fitting of the model for the COVID-19 outbreak: Italy and India

We consider three scenarios to fit and estimate parameters for our model. First, for Italy, where the epidemic was controlled and currently have a resurgence with increasing number of cases. Second, for India, which is observing significant number of cases in recent times. Third, for Victoria, Australia, where a second reemergence is controlled with strict intervention.

For Italy, we use data from the official source (the Department of Civil Protection - Presidency of the Council of Ministers) about the evolution of the epidemic in Italy from 2020-02-24 to 2020-08-27. We convert this data to fraction of population by taking total population data from The World Bank Group (about 60297396, in 2019). The estimated parameter values are based on the data about the number of currently infected individuals that can be observed and roughly corresponding to (*Q*(*t*) + *H*(*t*) + *C*(*t*)), and the number of recovered individuals that can be observed and roughly corresponding to 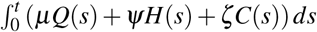. To avoid the pitfalls described by authors in^25^, we do not fit the model to cumulative number of cases or cumulative number of deaths - however, we present them for comparison.

The ordinary differential equation (ODE) system was solved using LSODA^26,27^. We use lmfit python package^28^ for non-linear least-squares and minimize of the sum of the squares of the errors using trust region reflective method and obtaining goodness of fit measure of *χ*^2^ = 5.3331*e*^−07^ for Italy. Confidence interval for the fit is shown in Figure 18a. The problem to minimize error is shown in following equation for fitting parameter set *p*:

**Figure 16.**
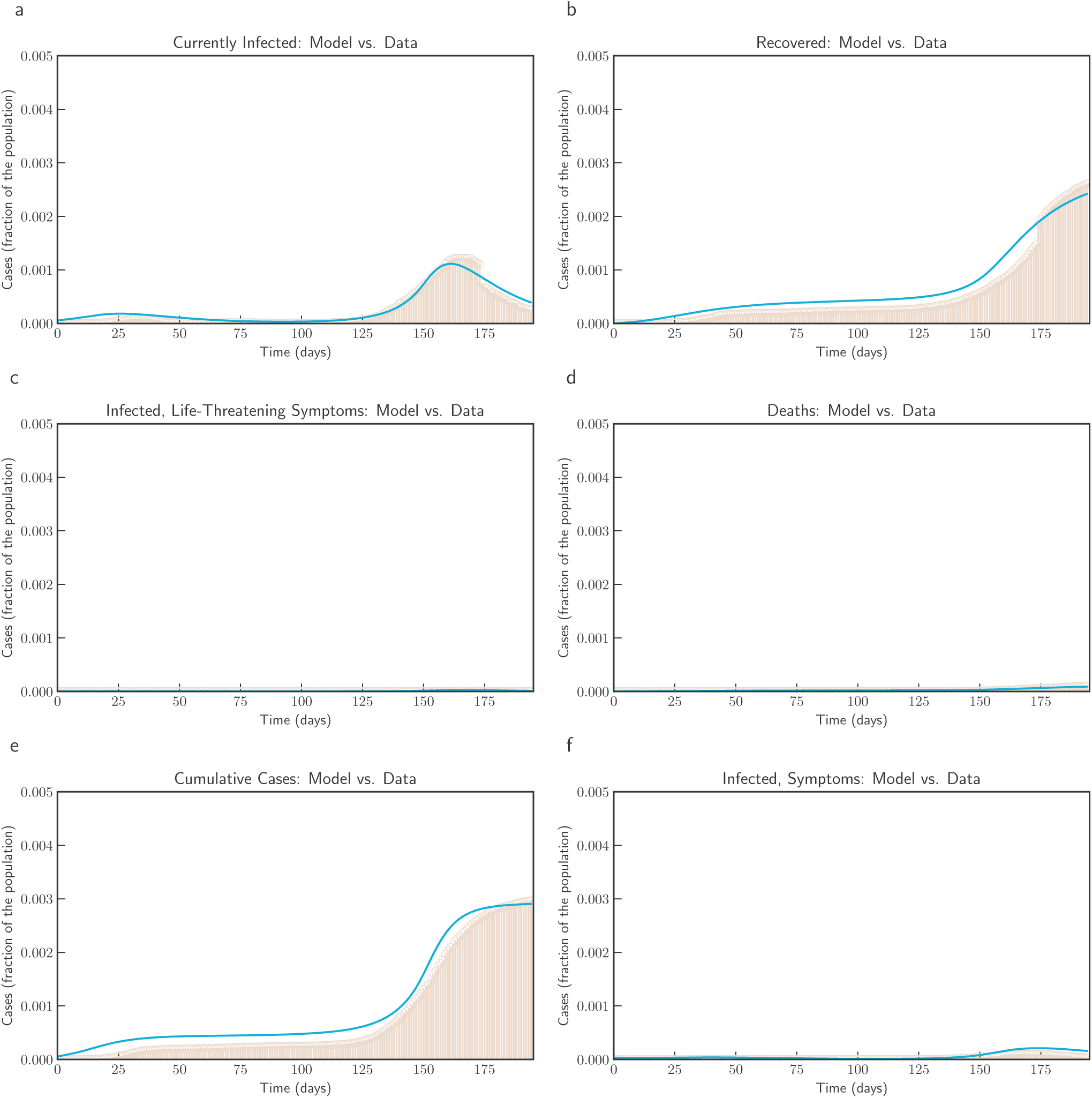
Model simulation compared to real data (Victoria, Australia) - Comparison between the official data (histogram) and the results with our model. Description of panels: **(a):** Number of currently active cases, (*Q*(*t*) + *H*(*t*) + *C*(*t*)), **(b):** number of reported recovered individuals. 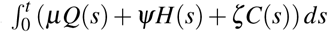, **(c):**number of reported infected with life-threatening symptoms, admitted to ICU, *C(t)*, **(d):** Number of deceased individuals *D(t)*, **(e):** Cumulative number of cases,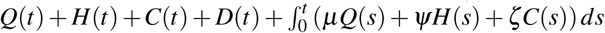 **(f):** number of reported infected with symptoms, who are hospitalized. *H(t)*

**Figure 17.**
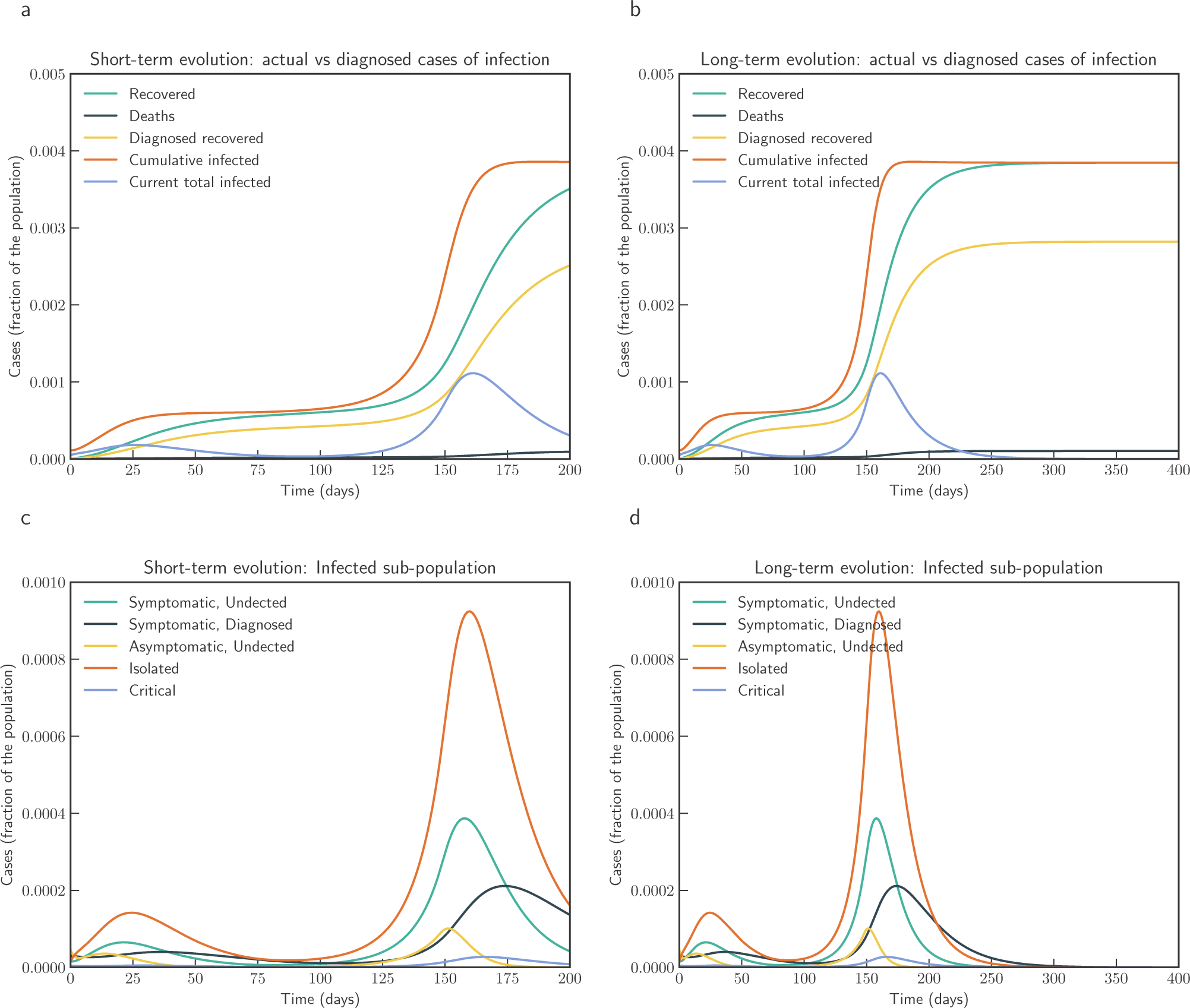
Model simulation compared to real data (Victoria, Australia) - Epidemic evolution predicted by the model based on the available data. Description of panels: **(a, c):** The short-term epidemic evolution obtained by reproducing the data trend with the model, **(b, d):** Long term epidemic evolution over 350 days. Plots refers to all cases of infection, both diagnosed and non-diagnosed, predicted by the model, although non-diagnosed cases are of course not counted in the data. Note that not all panels are in the same scale.

**Figure 18.**
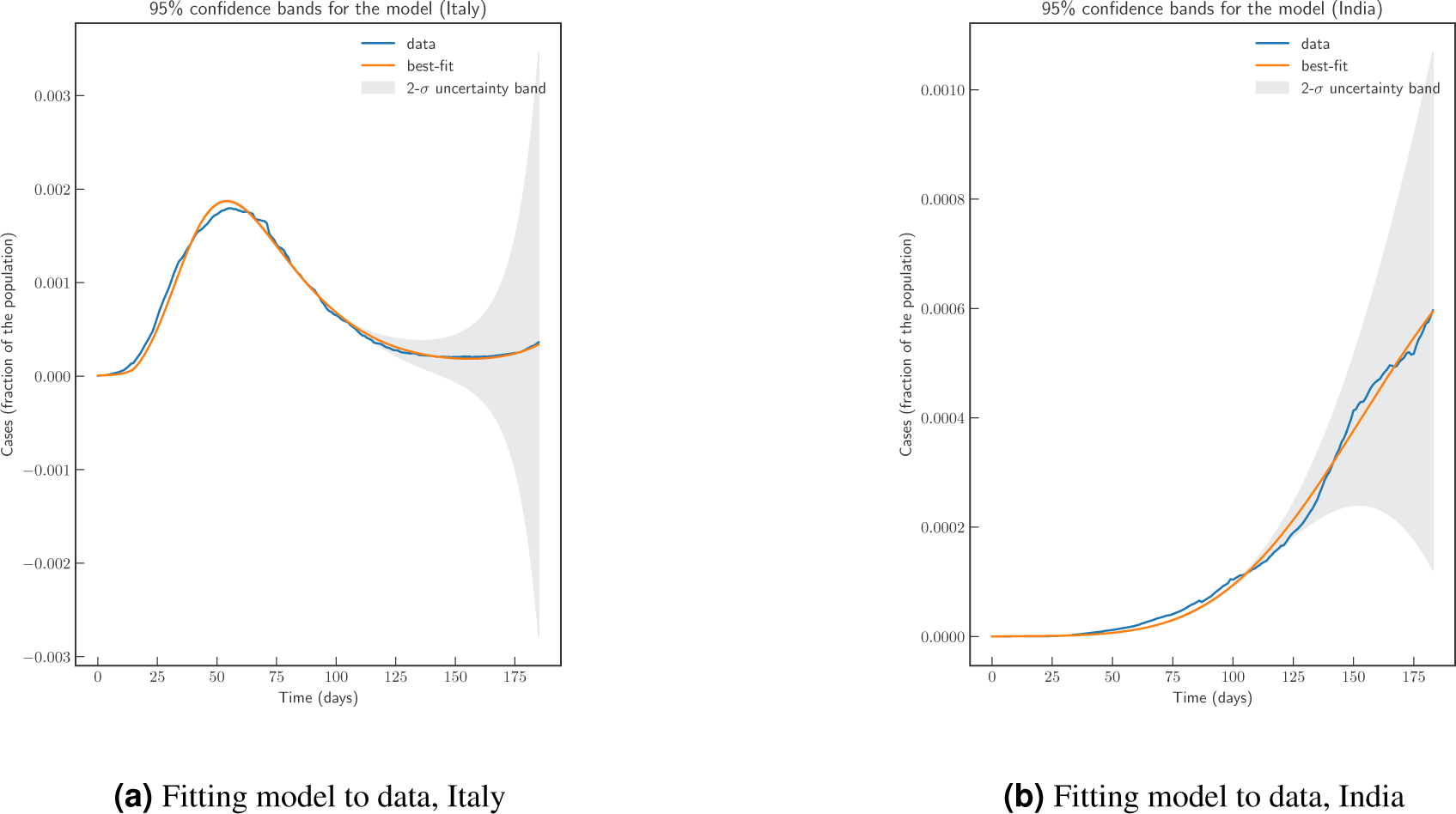
Parameter estimation and fitting model to actual data for Italy and India using lmfit. The plot show actual data, best fit by minimizing sum of the squares of the errors, and 2-*σ* uncertainty band

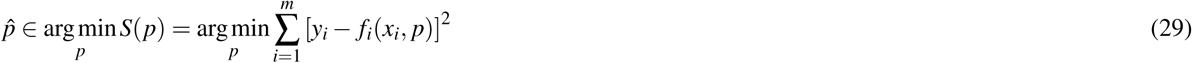

where *y*_*i*_ are observations and *f*_*i*_ is the model output.

life-threatening symptoms, admitted to ICU, *C*(*t*), **(d):** Number of deceased individuals *D*(*t*), **(e):** Cumulative number of cases,*Q*(*t*) + *H*(*t*) + *C*(*t*) + *D*(*t*) + J ^*t*^ (*µQ*(*s*) + *ψH*(*s*) + *ζC*(*s*)) *ds* **(f):** number of reported infected with symptoms, who are hospitalized. *H*(*t*)

For India (population 1366417754, in 2019), we use data from Johns Hopkins University Center for Systems Science and Engineering (JHU CSSE). We use data from 2020-03-02 to 2020-09-16. The estimated parameter values are based on the data about the number of currently infected individuals that can be observed and roughly corresponding to (*Q*(*t*) + *H*(*t*) + *C*(*t*)). In this case we obtain a goodness of fit measure of *χ*^2^ = 4.4811*e*^−08^ and confidence interval for the fit is shown in Figure 18b. Parameters *β*_0_, *β*_min_, *β*_*new*_, *t*_0_, *t*_1_, *r, u, ε*_0_, *ε*_max_, *t*_2_ and *s* have all been estimated from fitting the model to data. It is important to note that contact rat *β* (*t*) and testing rate *ε*(*t*) has been modeled as time variant functions.

For the state of Victoria, Australia, we use data from Covid19data.com.au, which is independent and voluntarily-run. We use data from 2020-02-27 to 2020-09-09. The estimated parameter values are based on the data about the number of currently infected individuals, and we obtain a goodness of fit measure of *χ*^2^ = 1.6701*e*^−06^.

### Extending the baseline model

In this section we consider various extension of the model and parameters.

### Scenario: Confinement

First extension is to model confinement - where a large proportion of susceptible population is placed under restricted movement, and thereby reducing the probability of contact. Such restriction has been modeled by introducing another compartment *L*. Individuals from *S* move to *L* at rate *u*(*t*). In many countries, confinement was spread over several weeks followed by a period of de-confinement. We model de-confinement by individuals moving from *L* to *S* at rate *p*(*t*). In specific we modify equation (9) as follows

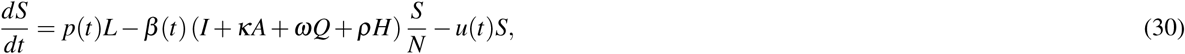

We also add following dynamics to the baseline model

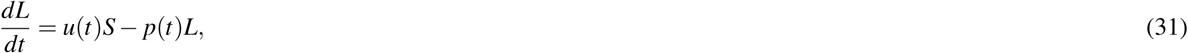

Thus, equations (1-8), equation (30), and (31) along with equation (10), defines the confinement - de-confinement scenario. A few remarks are in place for the time dependent functions *u*(*t*) and *p*(*t*).

### Scenario: Loss of immunity

To consider the possibility of loosing acquired immunity over time and having a reinfection, we modify equation (9) and add a term +*χR* and modify equation (8) by adding term −*χR* to obtain:

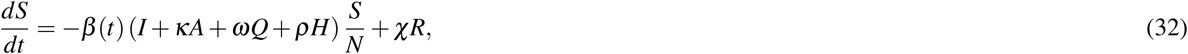

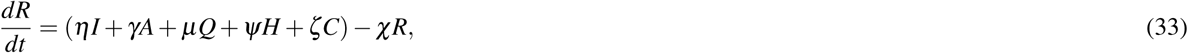

### Scenario: Mass Vaccination

In this scenario, we assume that the susceptible individuals are vaccinated at rate *ξ* (*t*). Initially *ξ* (*t*) can be small and as more vaccines are produced at a larger scale, the waiting time to receive vaccine reduces (see equation (34)).

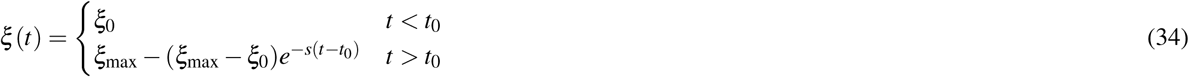

We also assume a vaccine efficiency parameter, and assume that vaccine does not confer immunity to all vaccine recipients, and hence a vaccinated individuals may become infected but at a lower rate than un-vaccinated. Hence, by (1 − *ϕ*) we denote vaccine efficiency. Thus, effective contact rate is multiplied by a scaling factor of *ϕ* 0 ≤ *ϕ* ≤ 1, where *ϕ*= 0 represents vaccine that offers 100% protection against infection.

The vaccinated individuals will be denoted with compartment *V*. With that, the augmented model can be represented with following system of equations, where we replace equation (1) with (35), equation (9) with (37), remaining equations (2)-(8) remains same and we add equations (36), and ((38):

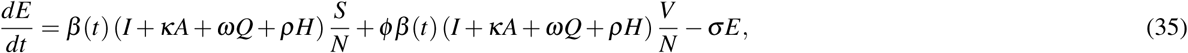

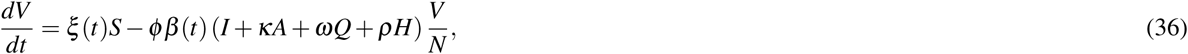

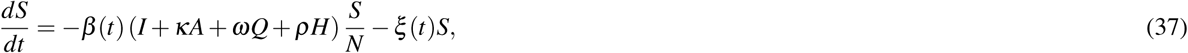

where,

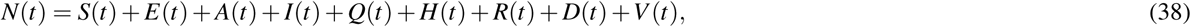

is the total population.

## Data Availability

All of the data are publicly available and has been extracted from https://github.com/pcm-dpc/COVID-19, from https://github.com/CSSEGISandData/COVID-19, and from Covid19data.com.au

https://github.com/CSSEGISandData/COVID-19

https://www.covid19data.com.au/

## Authors’ contributions

S. K. G.: Conceptualization, Data Collection, Software, Visualization, Writing - Original Draft, Writing - Review and Editing. S. G.: Conceptualization, Writing - Original Draft, Writing - Review and Editing. All authors reviewed the manuscript.

## Data availability

All of the data are publicly available and were extracted from https://github.com/pcm-dpc/COVID-19, and from https://github.com/CSSEGISandData/COVID-19.

## Code availability

Python codes were used for model implementation. All source codes can be accessed from https://github.com/subhaskghosh/model_covid_19

## Competing Interests statement

The authors have no competing interests.

## Notes

### Competing Interest Statement

The authors have declared no competing interest.

### Funding Statement

The author(s) received no specific funding for this work.

